# Monocyte, Neutrophil and Whole Blood Transcriptome Dynamics Following Ischemic Stroke

**DOI:** 10.1101/2022.03.03.22271866

**Authors:** Paulina Carmona-Mora, Bodie Knepp, Glen C Jickling, Xinhua Zhan, Marisa Hakoupian, Heather Hull, Noor Alomar, Hajar Amini, Frank R Sharp, Boryana Stamova, Bradley P Ander

## Abstract

**Background:** After ischemic stroke (IS), peripheral leukocytes infiltrate the damaged region and modulate the response to injury. Peripheral blood cells display distinctive gene expression signatures post IS and these transcriptional programs reflect changes in immune responses to IS. Dissecting the temporal dynamics of gene expression after IS improves our understanding of immune and clotting responses at the molecular and cellular level that are involved in acute brain injury and may assist with time-targeted, cell-specific therapy.

**Methods:** The transcriptomic profiles from peripheral monocytes, neutrophils, and whole blood from 38 ischemic stroke patients and 18 controls were analyzed with RNAseq as a function of time and etiology after stroke. Differential expression analyses were performed at 0-24 h, 24-48 h, and >48 h following stroke.

**Results:** Unique patterns of temporal gene expression and pathways were distinguished for monocytes, neutrophils and whole blood with enrichment of interleukin signaling pathways for different timepoints and stroke etiologies. Compared to control subjects, gene expression was generally up-regulated in neutrophils and generally down- regulated in monocytes over all times for cardioembolic, large vessel and small vessel strokes. Self-Organizing Maps identified gene clusters with similar trajectories of gene expression over time for different stroke causes and sample types. Weighted Gene Co-expression Network Analyses identified modules of co-expressed genes that significantly varied with time after stroke and included hub genes of immunoglobulin genes in whole blood.

**Conclusions:** Altogether, the identified genes and pathways are critical for understanding how the immune and clotting systems change over time after stroke. This study identifies potential time- and cell-specific biomarkers and treatment targets.

## Background

Ischemic stroke (IS) is one of the leading causes of death and disability in the world. Brain injury follows arterial occlusions in large or small cerebral vessels. These may arise due to several different causes that ultimately deprive the tissue of necessary oxygen and glucose [1]. Effective treatments are limited to short time windows and access to stroke centers. Early diagnosis is paramount for best outcomes. Current differential diagnosis usually requires advanced brain imaging. Therefore, there is a need for tests that utilize reliable and accurate molecular biomarkers from blood.

The immune and clotting systems play critical roles in the injury and recovery from stroke. After IS, peripheral leukocytes, including monocytes and neutrophils, infiltrate the injured area, mediating the immune response that causes inflammation and subsequent resolution and repair [2, 3]. Monocytes are composed of different subsets: classical (pro-inflammatory CD14^++^ CD16^−^), intermediate (CD14^++^ CD16^+^), and nonclassical (anti-inflammatory CD14^+^ CD16^++^) [4]. In addition, monocytes and neutrophils undergo polarization after IS: activated M1 monocytes or monocyte-derived macrophages and N1 neutrophils that are related to the inflammatory response can polarize to M2 or N2 phenotypes that are associated with the resolution and regenerative phase [5–8].

In models of experimental IS, neutrophils increase in the brain after 3 h and reach peak levels in the first 24 h [9, 10]. Monocytes slowly infiltrate the injury, peaking at day 3 or later after experimental stroke [2, 11]. Neutrophil levels in the brain return to near normal by a week after stroke, while monocyte increases persist for over a month [2, 9]. The prolonged monocyte/macrophage presence is likely indicative of ongoing peripheral leukocyte interaction with the injured brain associated with recovery phases. Analyzing peripheral leukocytes after stroke represents a feasible proxy to study the cellular and pathological changes that occur in response to the brain parenchyma injury. An increase of circulating neutrophils occurs promptly after stroke, and altered ratios of peripheral leukocytes (including neutrophils and monocytes) are indicators of outcome [12–17]. Successful intervention strategies in the acute and subacute phases of stroke may be improved when the pathological role of specific leukocyte types at different times is considered.

Transcriptional changes are detected promptly after IS in peripheral blood cells, showing how dynamic changes in gene expression can be revealed even in the acute phase of stroke. This results in distinct signatures depending on the cell type and stroke etiology [18–22]. Peripheral monocytes and neutrophils have been shown to be major cell types that display a transcriptomic response within the first 24 h after stroke [23]. In this study, the transcriptomic profiles from peripheral monocytes and neutrophils and whole blood were analyzed as a function of time and of different etiologies. Different analytical approaches (differential expression, self-organizing maps, and Weighted Gene Co-expression Network Analysis (WGCNA [24]) enabled the identification of genes that change expression following acute stroke. Though changes of gene expression in whole blood have been described within 0 to 24 hours following ischemic stroke using microarrays [23, 25], this is the first study to analyze the transcriptional profiles of monocytes, neutrophils and whole blood with RNA-seq at times ranging from 0 to >48 hours. The focus on the response over time in different cell types is crucial for the eventual development of diagnostic biomarkers and cell- and time-tailored treatments.

## Methods

### Subjects

Thirty-eight ischemic stroke (IS) patients and 18 vascular risk factor control (VRFC) subjects were recruited at the University of California at Davis Medical Center under a study protocol reviewed and approved by the Institutional Review Board (IRB- ID 248994-41). The study adheres to federal and state regulations for protection of human research subjects, The Common Rule, Belmont Report, and Institutional policies and procedures. Written informed consent was obtained from all participants or a legally authorized representative.

The criteria for recruitment are detailed in our previous study [18]. Briefly, IS diagnoses (cardioembolic - CE, large vessel - LV, and small vessel/ lacunar - SV) were confirmed by two independent neurologists based on history, exam, brain CT or MRI and other testing. The exclusion criteria were: anticoagulation therapy (using coumadin, heparin or any NOACs), immunosuppressive therapy, current or recent (two weeks) infection, and hematological malignancies. Vascular risk factor control (VRFC) subjects had no history of stroke, myocardial infarction, or peripheral vascular disease and they were recruited based on the presence of vascular risk factors including hypertension, hypercholesterolemia, and/or type 2 diabetes.

Whole blood for RNA analysis was drawn directly into PAXgene RNA stabilizing tubes for subsequent batch isolation. Blood for immune cell populations was collected in citrate tubes for immunomagnetic isolation by RoboSep (StemCell Technologies, Inc.). Monocytes were positively selected using antibodies to CD14 to a purity of >93% and neutrophils were enriched by negative selection to a purity of >99% as previously validated by flow cytometry [18].

### RNA-Sequencing and differential gene expression analyses

RNA isolation and cDNA library preparation were performed as previously described [18]. In summary, total RNA was extracted from isolated monocytes and neutrophils using the Zymo Direct-zol RNA mini-prep kit (Zymo Research) according to the manufacturer’s protocol. This was followed by treatment with DNase (QIAgen). Total RNA from whole blood samples was extracted using QIAcube with PAXgene Blood miRNA Kit (QIAgen). Ribosomal RNA and globin transcripts were depleted using InDA- C (aka AnyDeplete) during library preparation by the NuGEN Ovation Universal RNA-Seq system (Tecan Genomics, Inc.). RNA sequencing yielded an average of 200 M ± 10 M 2x150 bp reads per sample. Raw data were processed to generate counts as previously described [18]. Partek Genomics Suite was used for differential expression with an analysis of covariance (ANCOVA) model with REML variance estimate using the model: Y *ijklmn* = *μ* + Diabetes *i* + Diagnosis *j* + Hypercholesterolemia *k* + Hypertension *l* + Time (h) + Diagnosis*Time Point (TP) *jm*+ *ε ijklmn*, where Y *ijklmn* represents the *n* ^th^ observation on the *i* ^th^ Diabetes, *j* ^th^ Diagnosis, *k* ^th^ Hypercholesterolemia, *l* ^th^ Hypertension, *m* ^th^ Time Point (TP), *μ* is the common effect for the whole experiment, and *ε* ijklmn represents the random error component [26].

To identify differentially expressed genes, subjects were split into time points (TPs) from stroke onset (TP1= 0-24 h; TP2= 24-48 h; and TP3= > 48 h; mean and SD for time (h) in every time point are available in Table 1). Vascular Risk Factor Control (VRFC) subjects were assigned time point zero (TP0). Contrasts included time after stroke, interaction of diagnosis x TP, and major risk factor categories (diabetes, hypercholesterolemia, and hypertension) with cutoffs for significance set to p < 0.02 and fold-change > |1.2| to create lists of genes. Fisher’s least significant difference (LSD) was used for individual contrasts [27].

**Table 1:**
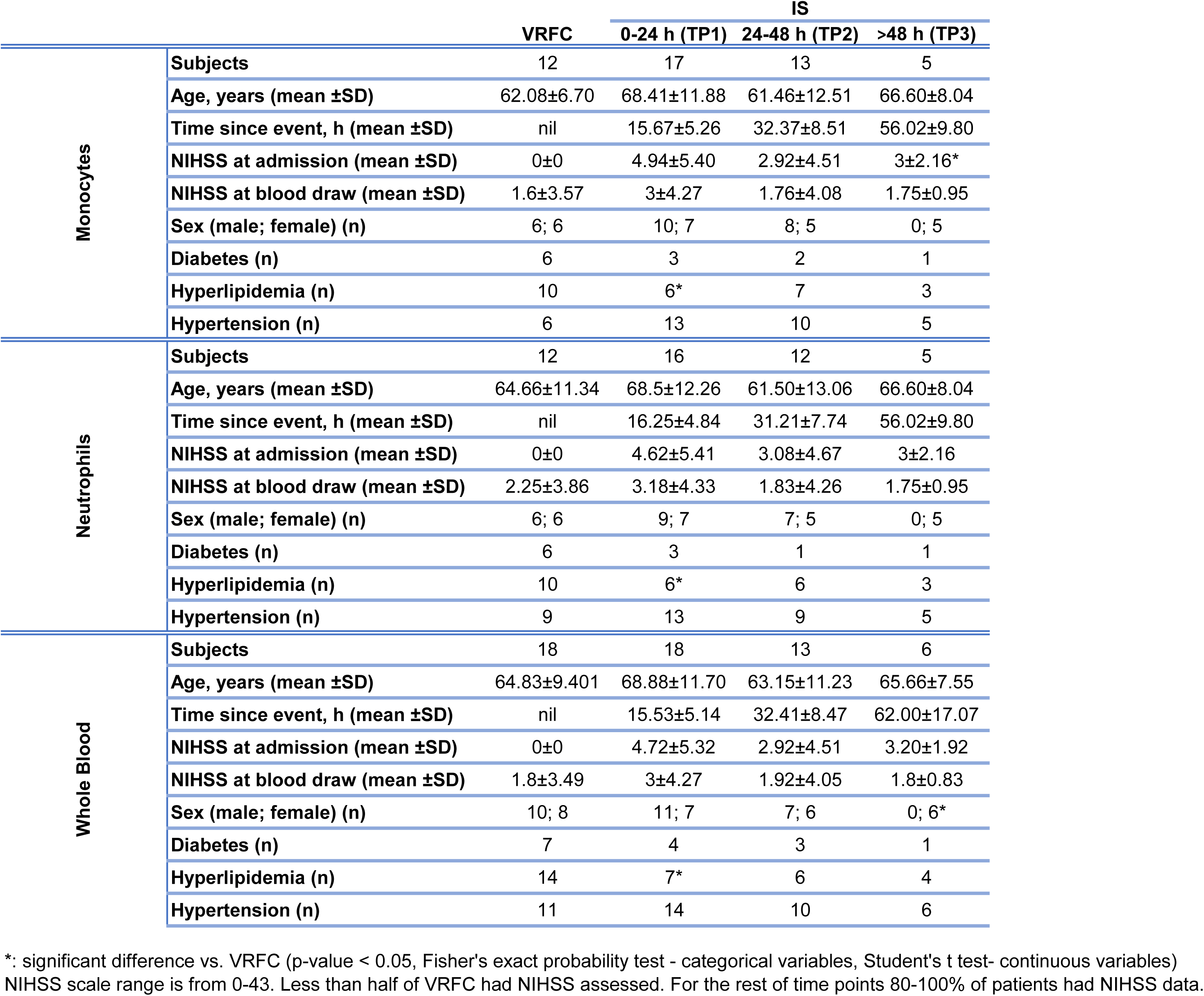
Subject demographics and relevant clinical characteristics in each of the time windows analyzed.

### Gene Clustering

Gene Expression Dynamics Inspector (GEDI) v2.1 [28] was used to create mosaic grids (tiles) of self-organizing maps for visualization of differentially expressed genes over time (https://github.com/midas-wyss/gedi). Two phases of training iteration were used (40 and 100) with linear initialization. Grid sizes were chosen depending on the total number of differentially expressed genes to analyze per sample type to keep a similar number of genes per tile in all mosaics (5x7, 7x8 and 4x6, for monocytes, neutrophils and whole blood samples respectively). Tiles corresponding to gene clusters of like-behaving genes were formed based on Pearson’s correlation. Tiles are composed of the same genes across time points, and mosaics for monocytes, neutrophils and whole blood have different tile composition.

Self-Organizing Maps (SOM) [29] were implemented in Partek Genomics Suite (alpha value set to 0.1, with random initialization, exponential decay function, gaussian neighborhood, and rectangular topology). This was done to examine trajectories of gene expression over time. The input for SOM consisted of differentially expressed genes that are present in at least two of the studied time-points. 500,000 training iterations were performed. Map height and width were set to 4×4 (16 profiles) in monocytes, neutrophils, and whole blood in the analyses irrespective of IS cause. For SOM in DEGs analyzed per IS etiology, map height and width were set as follows: 2×2 (CE), 2×3 (LV) and 2×2 (SV) in monocytes; 2×2 (CE), 3×3 (LV) and 2×3 (SV) in neutrophils; 2×3 (CE), 2×2 (LV) and 2×2 (SV) in whole blood (Supplemental Table 5). Differentially expressed gene expression was standardized by shifting to a mean of 0 (standard deviation (SD) of 1). Profiles were summarized and represented with ± 1SD. Profiles with similar dynamics were merged (based on similar directionality in every time point) for gene ontology analyses and visualization.

### Cell-specific markers

The presence or enrichment (p value < 0.05 for significant enrichment) of gene lists with blood cell type-specific genes was assessed by comparing to previously described blood cell type-specific genes [30, 31]. Enrichment analyses were performed using hypergeometric probability testing (R function *phyper*).

### Pathway and gene ontology analyses

Pathway enrichment analyses were performed using Ingenuity Pathway Analysis (IPA, Ingenuity Systems®, QIAgen). For input, differentially expressed genes and their fold-changes from every time point and sample type, with a p < 0.05 and fold-change > |1.2| were used. Pathways and predicted upstream regulators (Ingenuity Upstream Regulator analysis in IPA, white paper, Ingenuity Systems®, QIAgen) with Fisher’s exact test p < 0.05 were considered statistically overrepresented, and those that also have a Benjamini–Hochberg False Discovery Rate (FDR) correction for multiple comparisons are indicated in the Figures. IPA also computes significant pathway activation or suppression (z ≥ 2.0 and z ≤ −2.0, respectively), by using the z-score – which is based on comparisons between input data and pathway patterns, causal relationships, and curated literature. Gene ontology (GO) enrichment was explored as implemented in Partek Genomics Suite in the Gene Set Analysis, using Fisher’s exact test and FDR correction for multiple comparisons, with significance set at p<0.05.

### Weighted Gene Co-Expression Network Construction and Analysis

Separate weighted gene co-expression networks were generated for isolated monocyte (MON network) and neutrophil (NEU network) data, as well as for whole blood (WB network). VRFC samples were excluded, and genes below a minimum of 40 counts in every sample were filtered out for MON and NEU, and below a total of 80 counts for WB. The MON network was generated using the 14,955 detected genes after filtering across 35 IS samples; the NEU network was generated using 13,921 genes across 31 IS samples; the WB network was generated using 15,360 genes across 37 IS samples. Data were imported into R and checked for missing or zero-variance counts using the function *goodSamplesGenes*.

Networks were generated with the Weighted Gene Co-Expression Network Analysis (WGCNA) package using a Pearson correlation to measure co-expression [24]. An approximate scale free topology was depicted by the data. Soft thresholding powers (β) of 14, 8, and 16 were selected for the MON, NEU, and WB networks (Supplemental Figure 5), respectively, to maximize strong correlations between genes while minimizing weak correlations [32]. A signed network was used to consider both positive and negative correlations [33]. The *cutreeDynamic* function (method = tree; deepsplit = 1; minimum module size = 50) was used to form modules due to its adaptability to complex dendrograms and ability to identify nested modules [33]. Hub genes were defined as the top 5% by interconnectivity and may represent genes with important regulatory or molecular signaling roles [34, 35].

### Module Association with Clinical Parameters

Module-parameter associations were determined using Kruskal-Wallis and Spearman Ranked Correlation tests in Partek Genomics Suite for categorical and continuous variables, respectively. Ranked statistical tests were utilized to minimize the impact of outliers. Parameters were associated with the module eigengene, or first principal component of expression of genes within a module. Continuous time since event and all clinical parameters (age, diabetes, hypercholesterolemia, hypertension, race, and sex) were examined on the complete datasets. A p-value < 0.05 was considered significant.

### Hub gene analyses

Functional modules were detected with HumanBase [36] in a tissue-specific manner, using all hub genes per sample type as input (monocytes and whole blood). Briefly, this tool is based on shared k-nearest-neighbors and the Louvain community-finding algorithm. Gene ontology terms in the results are considered significant based on a corrected P-value < 0.05 (one-sided Fisher’s exact tests with a Benjamini– Hochberg FDR correction for multiple comparison).

## Results

### Cohort demographics

Individuals were binned into time points (TPs) from stroke onset (TP1= 0-24 h, average ∼15 h; TP2= 24-48 h, average ∼32 h; and TP3= >48 h, average ∼56 h), and Vascular Risk Factor Controls (VRFC) were assigned TP0 (Table 1). The cohort demographics and clinical characteristics per time point are presented in Table 1, as well as the total number of subjects per subgroup. The statistical analysis of variables showed that age, diabetes and hypertension were not significantly different (p < 0.05) between TPs and controls in any of the sample types (monocytes, neutrophils or whole blood). NIHSS at admission was significantly higher in subjects for samples at TP3 when comparing to VRFC in monocytes. Hyperlipidemia was significantly different for TP1 in all sample types when comparing to VRFC, while sex is significantly different in whole blood at TP3. In this cohort, 5 out of 38 IS patients received thrombolytic therapy (recombinant tissue plasminogen activator, rtPA) within 4.5h of their stroke. None of the IS cases developed hemorrhagic transformation.

### Gene expression dynamics in the acute and subacute post-stroke phase

The dynamic changes in differential gene expression across different time windows were assessed in monocytes (M, Supplemental Tables 1A-C), neutrophils (N, Supplemental Tables 1 D-F) and whole blood (WB, Supplemental Tables 1G-I) using a significance cut off P<0.02 (Figure 1). In monocytes there were 290, 645 and 352 DEGs at 0-24 h (TP1), 24-48 h (TP2) and >48 h (TP3) compared to controls, respectively (Figure 1). In neutrophils, 508 (TP1), 745 (TP2) and 547 (TP3) genes were differentially expressed compared to controls. A total of 610 (TP1), 260 (TP2) and 227 (TP3) DEGs were identified in whole blood compared to controls (Figure 1 and Supplemental Table 1). Across the time points, the number of up- or down-regulated DEGs in each sample type showed distinct and dynamic trends. For example, in monocytes most DEGs are down-regulated, with the most at TP2 (Figure 1A, right panel). Conversely, in neutrophils most genes are up-regulated, peaking at TP2 (Figure 1B, right panel). In whole blood, the highest up-regulation occurs within the first 24 h (Figure 1C, right panel). Biotypes of DEGs were relatively similar in monocytes and neutrophils, while in whole blood more lncRNAs and T-cell receptor genes were detected at >48 h post-stroke (Supplemental Figure 1).

**Figure 1.**
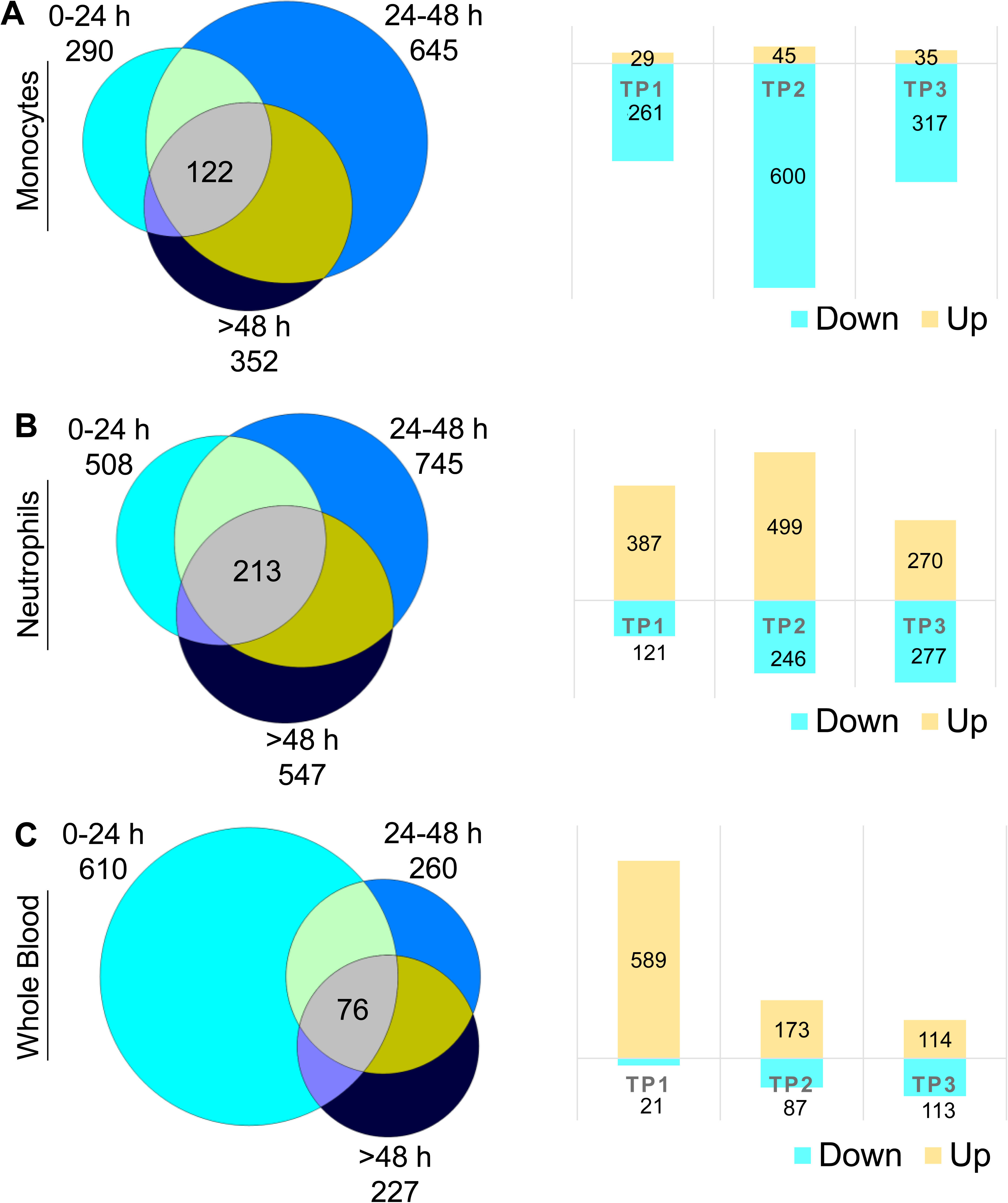
Differential expression across time points. Differentially expressed genes (DEGs) across time points versus VRFC (0-24 h (TP1), 24-48 h (TP2), and >48 h (TP3)) in monocytes (A), neutrophils (B) and whole blood (C). Venn diagrams of the numbers of DEGs at each time point are shown on the left, and on the right, the corresponding bar plots are shown for the numbers of up- and down-regulated DEGs found per time point.

Pathway enrichment analyses also show distinctive shifts in molecular function and cellular roles of the DEGs over time per sample type. Most of the overrepresented pathways in monocytes are shared across at least two TPs (Figure 2A) (average of 88.8% shared pathways) and most of these pathways are predicted to be significantly suppressed (Z ≤ -2.0), including chemokine, IL-8 signaling and production of NO and ROS in monocytes/macrophages (Supplemental Table 2A-C and Figure 2B and 2C). The only enriched pathways with predicted activation are PTEN signaling (at 24-48 h and >48 h) and RhoGDI signaling (at 24-48 h) (Supplemental Table 2B-C). Calcium signaling, CCR3, chemokine, CREB, and CXCR4 signaling were the top enriched pathways in monocytes at 0-24h (Figure 2C). Actin, B cell receptor, ephrin, Erb and ERK/MAK signaling were the top pathways over-represented in monocytes at 24-48 h (Figure 2C). Similar pathways were regulated in monocytes at times >48 h in addition to estrogen, FLT3, GMCSF, GNRH and HIF signaling (Figure 2C).

**Figure 2.**
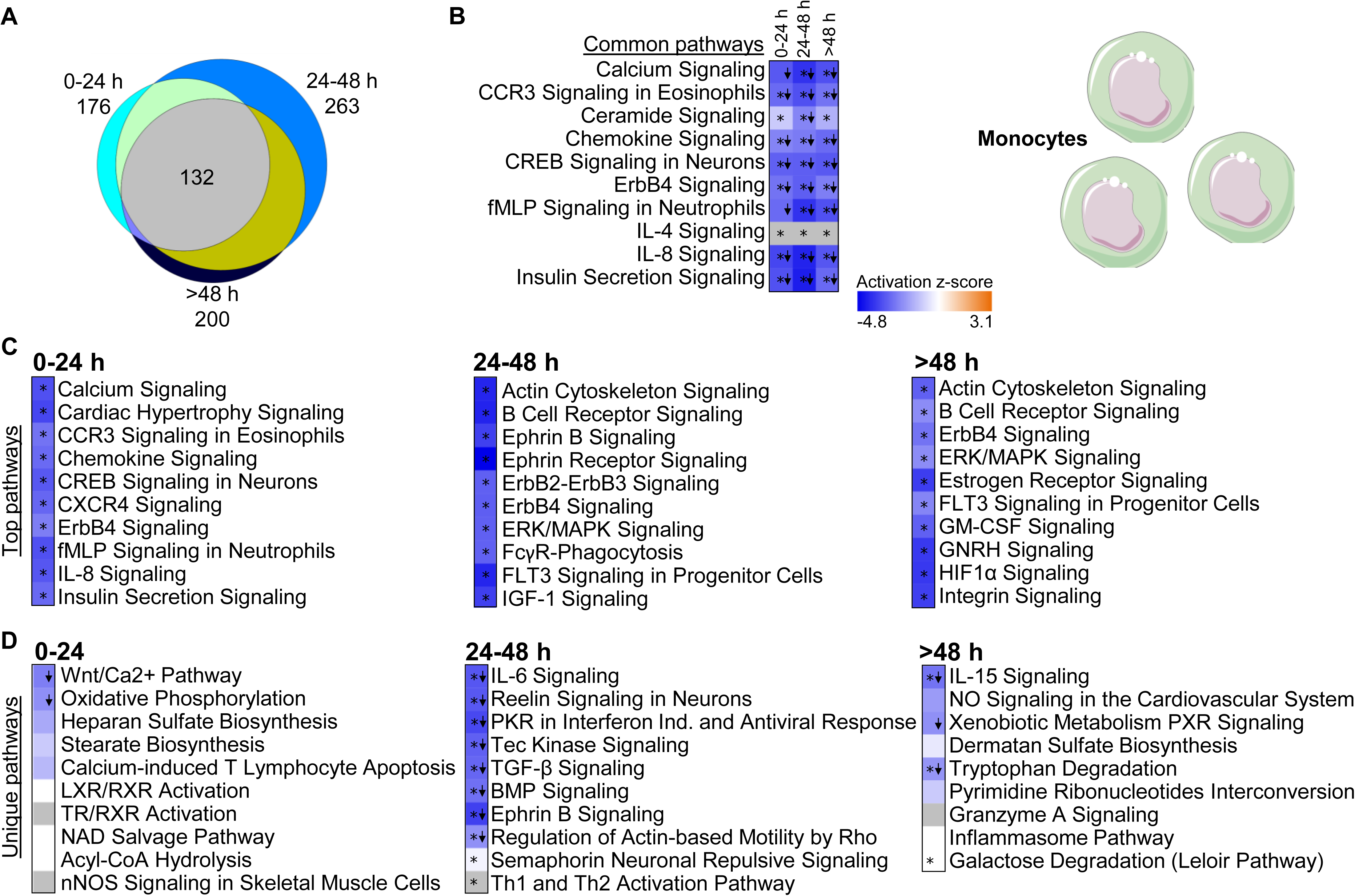
Enriched pathways in monocytes. (A) Venn diagram represents all enriched pathways in monocytes at 0-24 h, 24-48 h, and >48 h (Fisher’s p-value<0.05). 132 pathways overlapped all three time points. (B) Top 10 pathways enriched and shared at all time points. (C) Top 10 over-represented pathways with predicted inhibition (z≤ −2, blue shades) in every time point. (D) Top 10 over-represented pathways unique to each time point (Fisher’s p-value<0.05). Down arrows indicate significant predicted inhibition of pathway (z≤ −2). Blue cells indicate negative z score, white cells indicate no direction can be predicted, and grey cells are those where no prediction can be performed. (*= Benjamini-Hochberg corrected p-value<0.05). FcγR-Phagocytosis: Fcγ receptor-mediated Phagocytosis in Macrophages and Monocytes; Ind, induction.

From the pathways exclusively regulated at a single TP, suppressed oxidative phosphorylation, suppressed Wnt/Ca^2+^, heparan sulfate and nNOS pathways top the list in the first 24 h (Figure 2D, left panel). Ephrin B, Tec kinase, reelin, TGF-β and IL-6 signaling are amongst the suppressed pathways at TP2 (Figure 2D, middle panel); while Tryptophan degradation and IL-15 signaling are suppressed at times over 48h (Figure 2D, right panel).

In neutrophils, fewer enriched pathways were shared across at least two TPs (Figure 3A, Supplemental Table 2D-F) (average of 62.5% of shared pathways in TPs pairwise comparison). There were increased numbers of pathways at TP2 and TP3 compared to TP1 (Figure 3A, C). EIF2 signaling and leukocyte extravasation were over- represented at all TPs (Figure 3B). CD40 and HIF pathways are over-represented at TP2 and TP3. Several interleukin pathways are enriched in the later TPs (Figure 3C and Supplemental Tables 2D-F). IL22 was expressed at TP2 and TP3 (Figure 3C). IL-1, 4, 7 and 8 were exclusively overrepresented in TP2; while IL-2, 10 and 15 pathways were only enriched after 48 h (Figure 3D).

**Figure 3.**
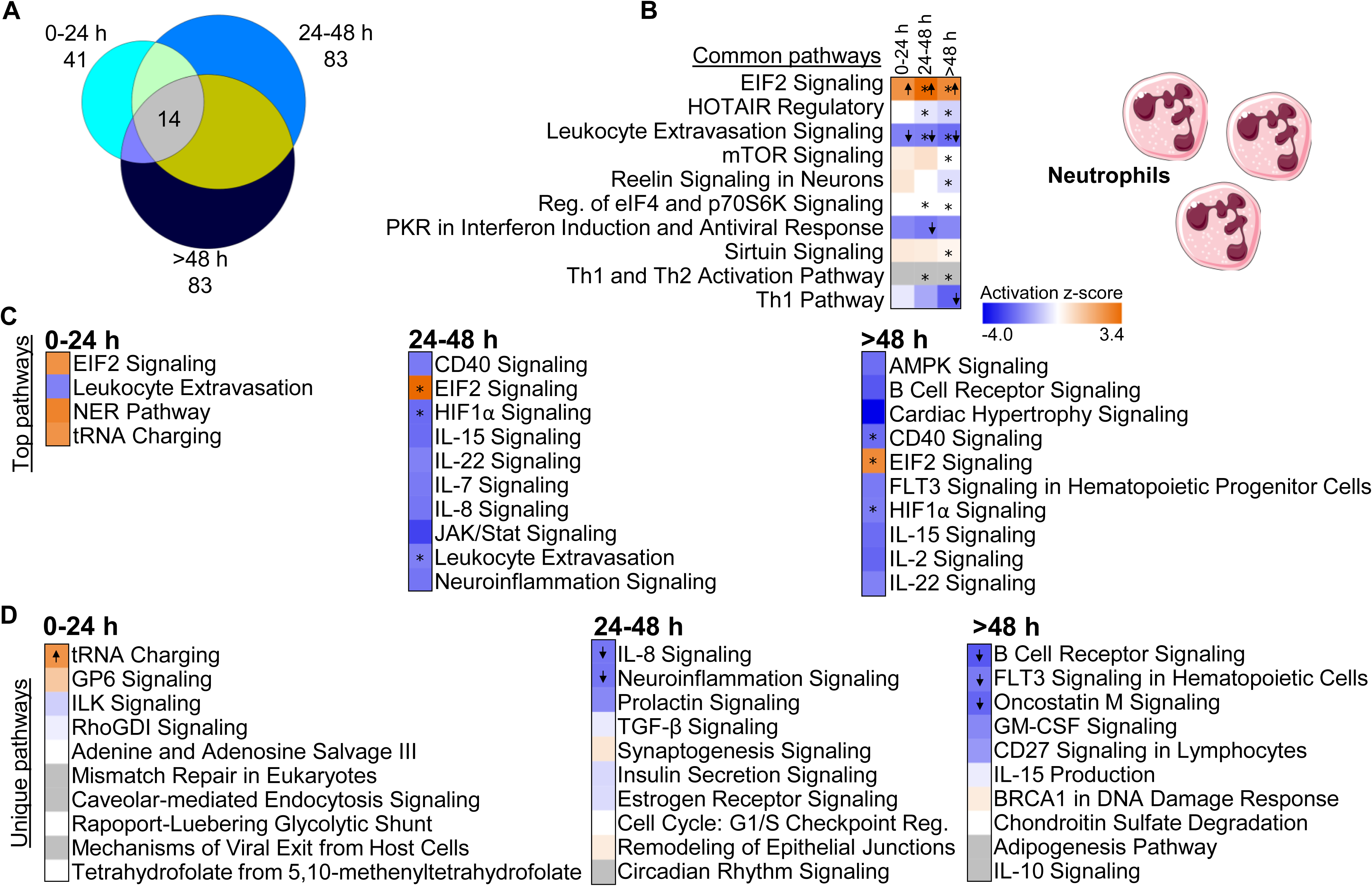
Enriched pathways in neutrophils. (A) Venn diagram represents all enriched pathways in neutrophils at 0-24 h, 24-48 h, and >48 h (Fisher’s p-value<0.05). 14 pathways overlapped all time points. (B) Top 10 common pathways enriched in all three time points. (C) Top over-represented pathways with predicted activation (z≥ 2, orange shades) or inhibition (z≤ −2, blue shades) at every time point. (D) Top 10 over- represented pathways unique for each time point (Fisher’s p-value<0.05). Down arrows indicate predicted significant inhibition (z≤ −2) and up arrows predicted significant activation (z≥ 2). Blue cells indicate negative z-score, and orange positive z-score. White cells indicate no direction can be predicted, and grey indicate that no prediction can be performed (*= Benjamini-Hochberg corrected p-value<0.05). Reg: regulation.

In contrast to the isolated cell samples, whole blood showed no shared pathways across all TPs (Figure 4A). Most pathways were enriched at only one TP in whole blood (Figure 4C and Supplemental Tables 2 G-I), and many of these were specific for 0-24 h including p53, AMPK and ATM signaling, and FXR/RXR activation (Figure 4C).

**Figure 4.**
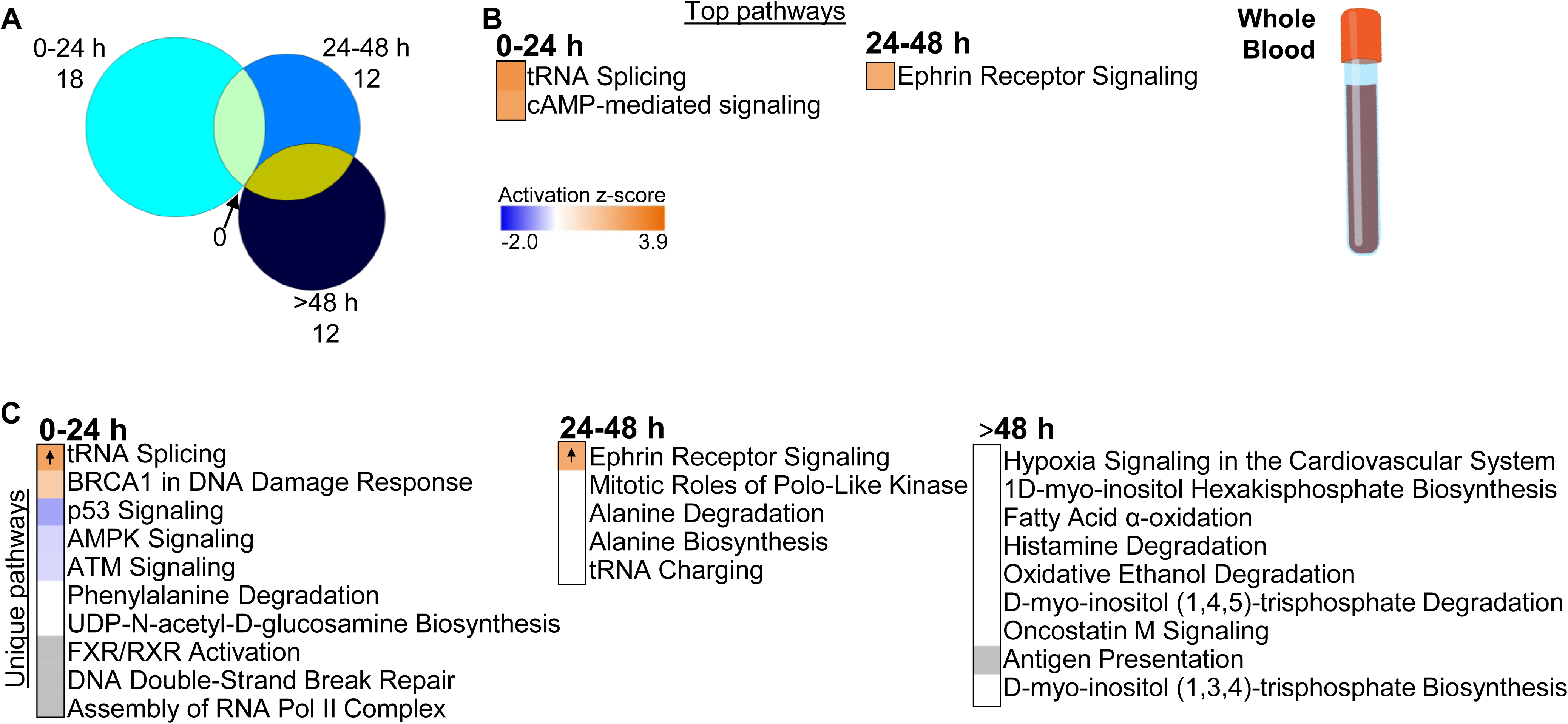
Enriched pathways in whole blood. (A) Venn diagram represents all enriched pathways at each of the time points (0-24 h, 24-48 h, >48 h; Fisher’s p- value<0.05). There was no overlap of pathways between the time points. (B) Top over- represented pathways with significant z-score (z ≥ 2, predicted activation) at 0-24 h and 24-48 h. No significant z-scores were predicted for pathways at >48 h. (C) Top over- represented pathways unique to each TP (Fisher’s p-value<0.05). Orange cells indicate positive z-score for the pathway, and blue indicate negative z-score. White bars indicate no direction can be predicted, and grey bars indicate no prediction can be performed. Up arrows indicate predicted significant activation (z ≥ 2).

Upstream regulators were computed to identify drivers of gene expression at specific TPs (Supplemental Table 3). Similar to what was seen with canonical pathway enrichment, there were more regulators shared across TPs for monocytes (Supplemental Figure 2, Supplemental Tables 3A-C) and neutrophils (Supplemental Figure 3; Supplemental Tables 3 D-F) compared to whole blood (Supplemental Figure 4; Supplemental Tables 3 G-I). Upstream regulators (both activators or suppressors) for monocytes included many molecules common to all time points:15-LOX, ACKR3, BTNL2, CSF2, ERG, ERK, filgrastim, IKZF1, lipopolysaccharide, Msx3, OGA, PCYT2, PGR, PLA2R1, Progesterone, Rhox5, TNF, U18666A and WAC (Supplemental Figure 2; Supplemental Tables 3A-C). Upstream regulators for neutrophils predicted at all time points included: calcitriol, CIP2A, ciprofloxacin, CSF3, dexamethasone, ESR1, filgrastim, FOXO3, FOXO4, interferon beta-1a, LARP1, MRTFB, MYCN, NUPR1, OSM, RPS15, RRP1B, torin1, TP53 and YAP1 (Supplemental Figure 3; Supplemental Tables 3 D-F).

In contrast, the vast majority of upstream regulators are unique for every TP in whole blood (Supplemental Figure 4; Supplemental Tables 3 G-I). In addition, there are more regulators in the first 24 h in whole blood (Supplemental Figure 4; Supplemental Tables 3 G-I), consistent with the proportion of whole blood DEGs seen at this time point.

### Identification of time dependent DEG clustering profiles

Tile mosaics based on self-organizing maps were constructed for DEGs with GEDI to allow an overall view of gene clusters across time by correlated expression.

These mosaics reveal the distinctive dynamics of gene expression changes across every time point in monocytes (Supplemental Table 4A), neutrophils (Supplemental Table 4B) and whole blood (Supplemental Table 4C) (Figure 5). Every tile clusters a specific group of DEGs, and the tile mosaics for every time point show how that group of genes changes over time (Figure 5 and Supplemental Table 4). Tracked at each coordinate, most tiles in the mosaic maps show evidence of dynamic changes at each time point (Figure 5, 0 to >48 h).

**Figure 5.**
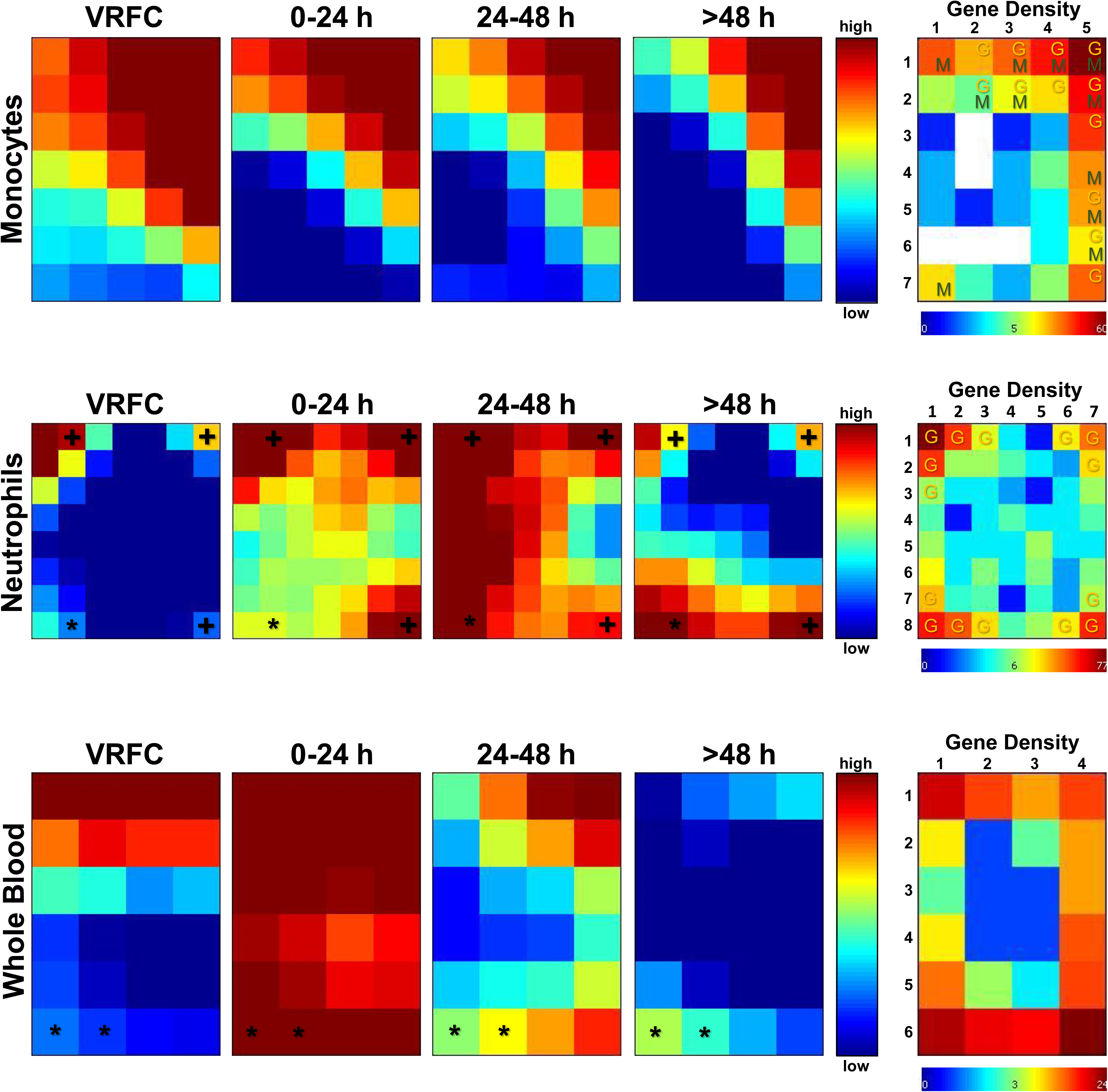
Dynamics of gene expression based on GEDI. Tile maps based on Self-Organizing Maps are shown for all time points for monocytes, neutrophils and whole blood. Each tile within a cell type represents the same set of genes across all 3 time points and VRFCs. Changes in color in a tile over the time points shows changes in expression of the member genes over time, while constant color over the time points shows more constant expression levels. The asterisk (*) indicates tiles where Immunoglobulin genes are present, and the plus (+) indicates the presence of interleukin receptor genes. On the right, gene density maps represent the number of DEGs per tile (genes clustered based on Pearson’s correlation). Tiles behaving similarly are placed in the same mosaic neighborhood. Letters on the tiles in the gene density map depict presence of specific cell markers in the DEGs in a specific tile (M: monocytes, G: granulocyte= neutrophils, basophils and eosinophils).

To understand the trajectory of the DEG clusters over time following stroke and to identify key genes per time window, we analyzed expression profiles using self-organizing maps (SOM) (Supplemental Table 5), where every cluster was plotted separately to obtain a profile. For each cell type including monocytes (Supplemental Table 5A), neutrophils (Supplemental Table 5B) and whole blood (Supplemental Table 5C) there were different patterns of gene expression that included increases over time, decreases over time, peaks at 24-48 h, and valleys at 24-48 h (Figure 6). Though comparable expression patterns between monocytes, neutrophils and whole blood are seen over time, these were associated with unique functions (GO) in each cell type (Figure 6, Supplemental Tables 6A-L). Examples include neutrophil degranulation which increases over time in whole blood, decreases of IL-1 secretion over time in monocytes, leukocyte migration peaking in neutrophils at 24 h, and a valley for regulation of platelet activation at 24-48 h in monocytes (Figure 6).

**Figure 6.**
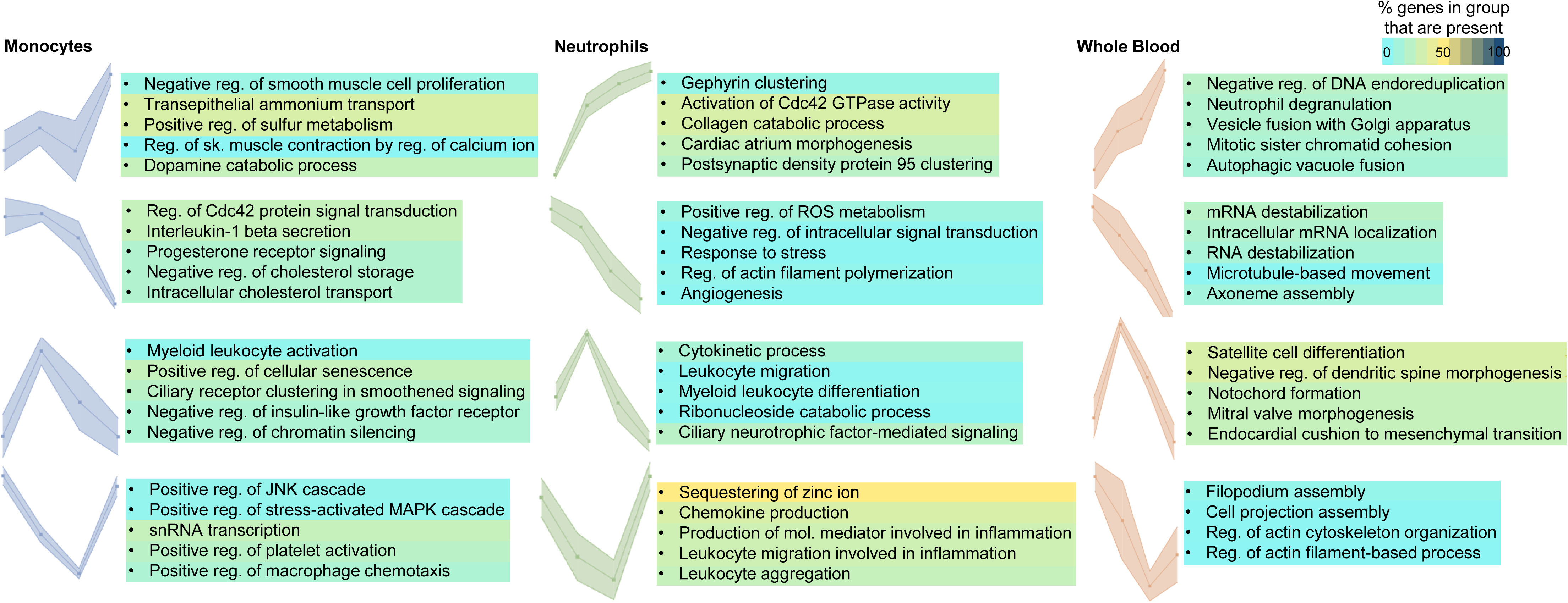
Distinctive profiles of gene expression over time after IS based on Self- Organizing Maps (SOM). DEGs profiles are shown across time points in monocytes, neutrophils and whole blood. Dots on each profile represent 0 (VRFC), 0-24 h, 24-48 h, and >48 h time points from left to right. Rows show similar expression profiles across sample types. The top significant Gene Ontology (GO) terms for biological processes are shown. Color corresponds to the percentage of genes in the GO group present in the profiles. Reg: regulation, sk: skeletal; mol: molecular.

### Time-dependent gene expression changes per stroke etiology

To identify expression changes associated with IS causes, the same model and criteria for DEGs were used, but time points were considered as 0-24 h and over 24 h (>24 h) post stroke in order to assure enough IS cases per etiology and time point. The cohort characteristics and analyses for the clinical variables after this re-stratification can be found in Supplemental Table 7. The cohort was split according to the main IS etiologies: cardioembolic (CE, Supplementary Tables 8 A, B, G, H, M, N), large vessel (LV, Supplementary Tables 8 C, D, I, J, O, P) and small vessel (SV, Supplementary Tables 8 E, F, K, L, Q, R).

More DEGs were identified in all IS causes vs. VRFC in the >24 h period than in the first 24 h for monocytes and neutrophils (Figure 7, Supplemental Table 8). In contrast, in whole blood the first day post-IS showed higher DEGs in CE and LV stroke, pointing to other critical cell types in the response within this time window (Figure 7 and Supplemental Table 8). Gene expression tended to be suppressed in monocytes following CE, LV and SV strokes at all time points, whereas gene expression was generally up-regulated in neutrophils in all stroke etiologies at all time points (Figure 7). Gene expression was increased in whole blood at 0-24 h in CV and LV strokes (Figure 7).

**Figure 7.**
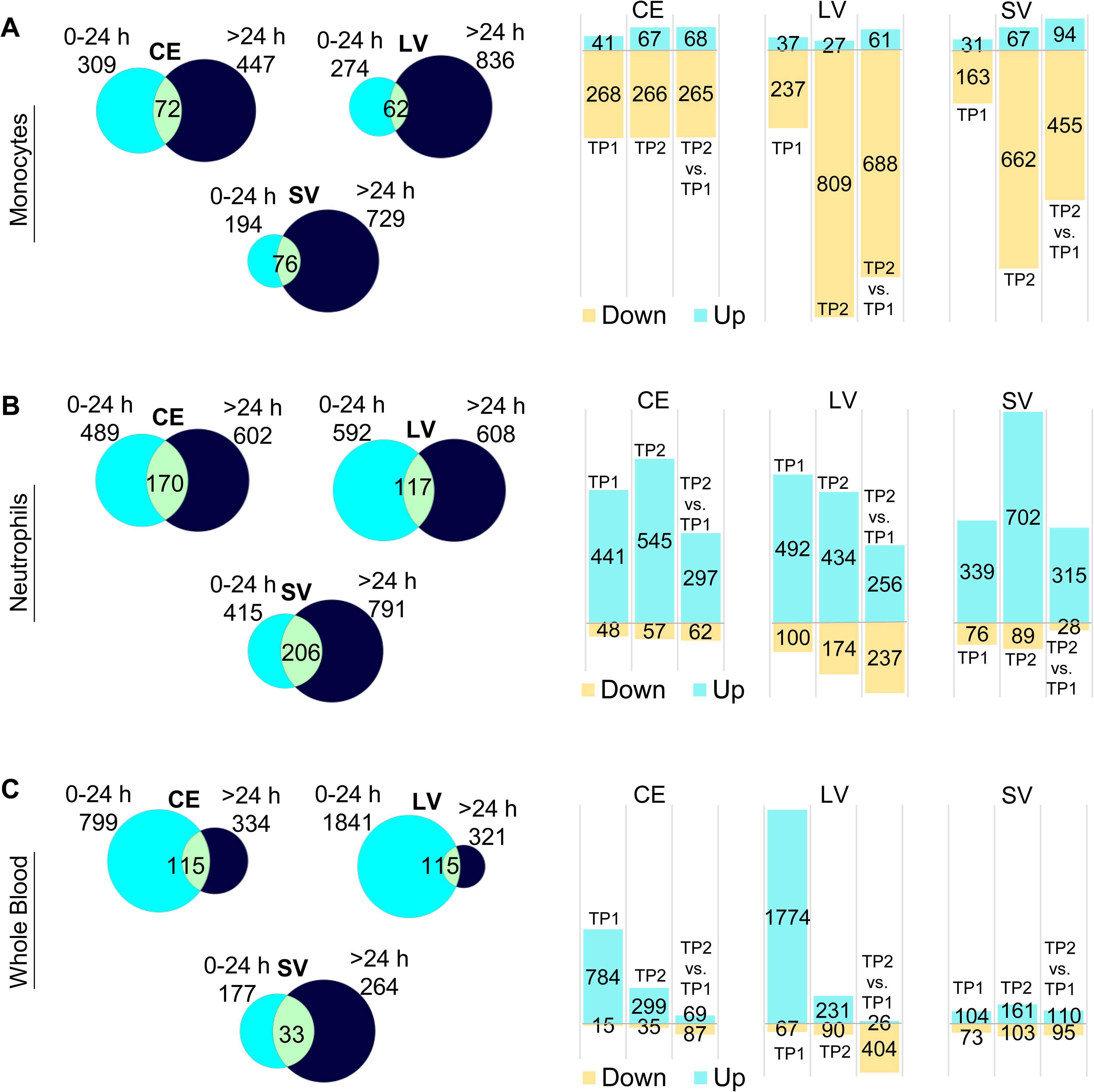
Differentially expressed genes (DEGs) in stroke etiologies. Venn diagrams of the numbers of DEGs at each time point versus VRFCs after cardioembolic (CE), large vessel (LV) and small vessel (SV) strokes in monocytes (A), neutrophils (B) and whole blood (C) (left panels). On the right are the corresponding bar plots for the numbers of up- and down-regulated DEGs found per time point and IS cause. (0-24 h (TP1), 24-48 h (TP2), and >48 h (TP3)).

Key DEG clusters per IS cause were identified using SOM for gene expression trajectories (Supplemental Table 9). Comparable expression trajectories over time are seen for CE, LV and SV strokes in monocytes (Supplemental Tables 9 A, B, C), neutrophils (Supplemental Tables 9 D, E, F) and whole blood (Supplemental Tables 9 G, H, I) (Figure 8), although not all trajectory types are consistently present in every IS cause and in every sample type (Figure 8).

**Figure 8.**
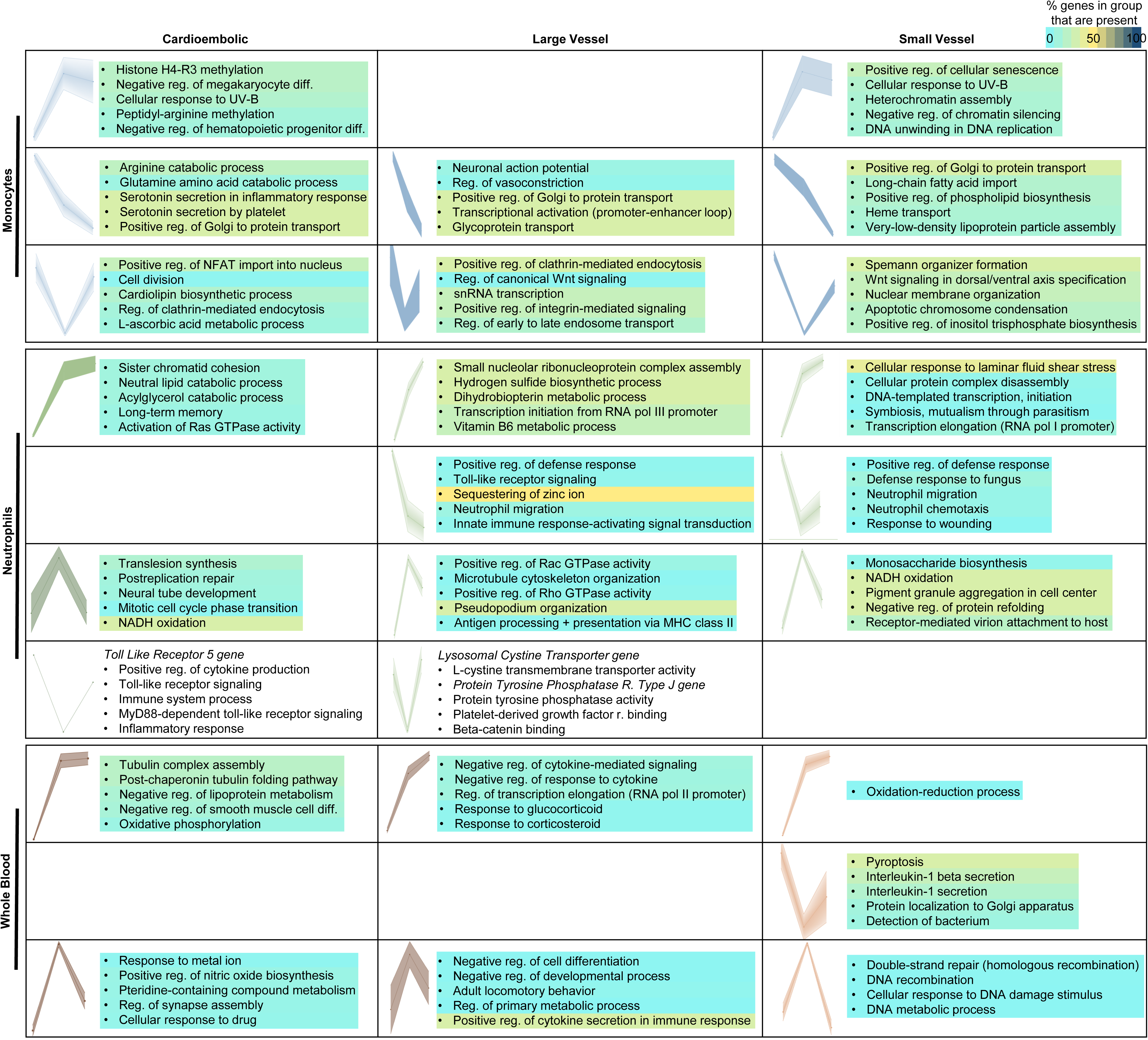
Distinctive gene expression trajectories over time for different causes of IS based upon Self-Organizing Maps (SOM). DEGs trajectories across time (0 h, 0-24 h, >24 h)) are shown for cardioembolic (CE), large vessel (LV) and small vessel (SV) causes of strokes in monocytes, neutrophils and whole blood. Dots on each profile represent every time point from left to right. Rows show similar expression profiles across stroke etiology in each sample type; empty cells indicate no matching profile was found in that sample type / etiology combination. The top enriched Gene Ontology (GO) terms for biological processes are shown for each profile. The color corresponds to the percentage of genes in the GO group present in the profiles. Reg: regulation; diff: differentiation; R: receptor.

GO enrichment revealed the associated functions for similar profile trends in every sample type, which tended to be unique for each IS etiology and cell type (Supplemental Tables 10 A-W; Figure 8). In neutrophils, for example, the GO terms neutrophil migration and positive regulation of defense response are enriched in the profile of genes that decrease expression throughout TPs in LV and SV strokes, while it is not present in CE (Figure 8). Interestingly, regulation of neutrophil migration is enriched in the profile of DEGs that consistently increase expression in LV stroke in the whole blood samples (Supplemental Table 10).

### Time-associated gene expression networks/modules in monocytes, neutrophils and whole blood

To analyze the time-dependent changes that occur after IS from a genome-wide perspective, we used Weighted Gene Co-expression Network Analysis (WGCNA). The gene expression counts for monocytes, neutrophils and whole blood samples from IS patients were used to generate three separate networks, and significant modules of co-expressed genes were identified in relation to time (as continuous variable in hours post-IS) (Figure 9A, Supplemental Table 11).

**Figure 9.**
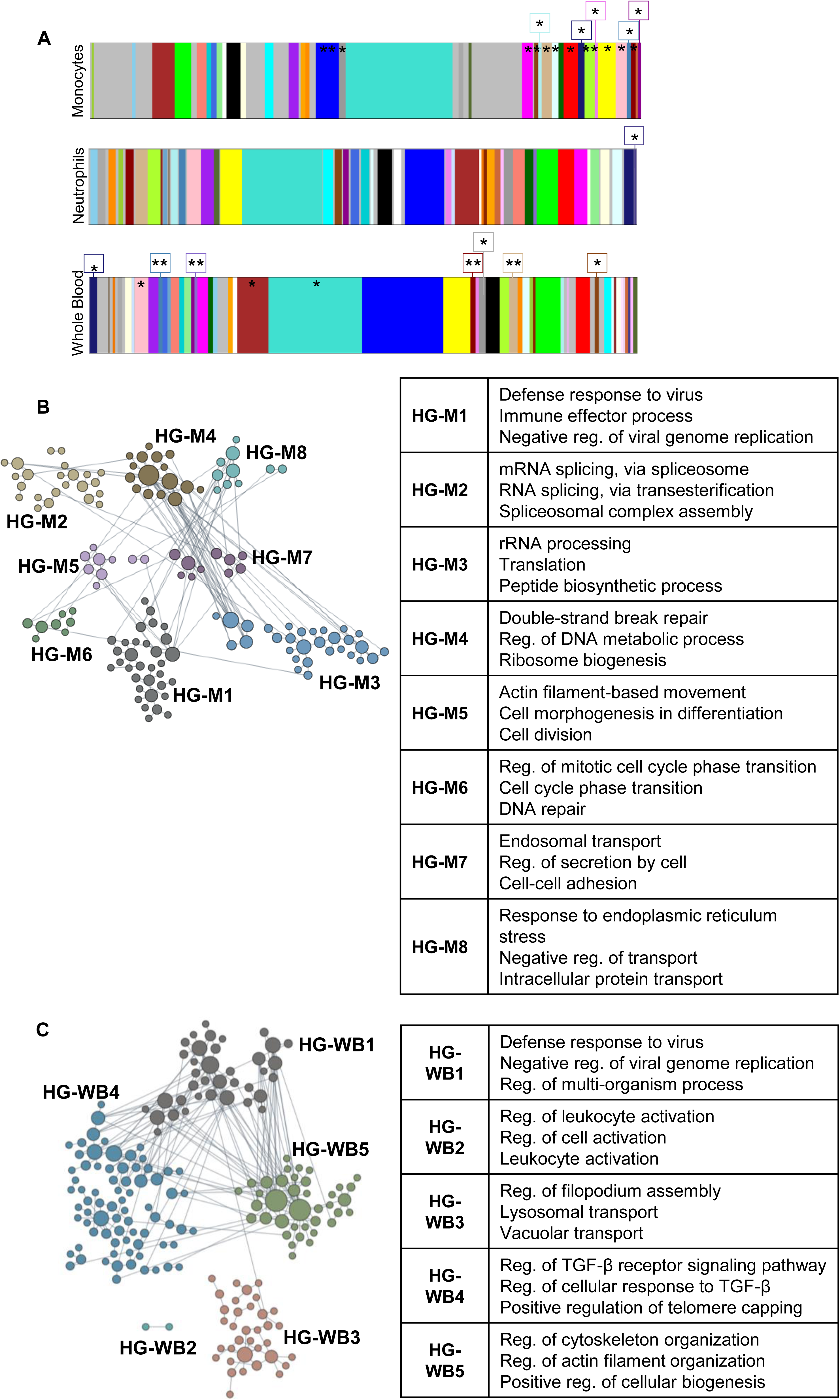
Co-expression modules of genes significant to time after stroke, hub genes and their functional clusters. A) All WGCNA-identified gene co-expression modules in monocytes, neutrophils and whole blood. Modules significant for time are labeled with an asterisk (*) and those additionally significant to another factor with two asterisks (**). Hub genes that varied as a function of time were clustered according to their functional organization (HG: hub gene module) in monocytes (HG-M) as shown in (B) (left panel), and in whole blood (HG-WB) in (C) (left panel). The top 3 most specific significant GO terms in each hub gene functional cluster are shown for monocytes in (B) (right panel) and for whole blood in (C) (right panel) (images modified from HumanBase). Reg: regulation.

For monocytes, 36 modules of co-expressed genes and a module of unassigned genes (MON-Grey) were identified within this cell type network. 16 monocyte modules were found to be significant for time: MON-DarkRed, MON-MidnightBlue, MON- GreenYellow, MON-Sienna3, MON-Red, MON-Pink, MON-LightCyan, MON-Violet, MON-Yellow, MON-DarkMagenta, MON-SteelBlue, MON-Blue, MON-Grey60, MON- Tan, MON-PaleTurquoise, and MON-Magenta (Figure 9A). The gene lists for each of these 16 modules are provided in Supplementary Table 11A. Of these, MON- GreenYellow and MON-Blue were also significantly related to diabetes. In neutrophils, 48 modules of co-expressed genes and a module of unassigned genes (NEU-Grey) were identified within the network (Figure 9A). One neutrophil module (NEU- DarkSlateBlue) was significant for its relationship to time. The gene list associated with this one neutrophil module is provided in Supplementary Table 11B.

Within the whole blood network, 51 modules of co-expressed genes and a module (WB-Grey) of unassigned genes were identified. Ten of these whole blood modules were significantly related to time: WB-Brown, WB-Grey60, WB-Tan, WB- Turquoise, WB-MidnightBlue, WB-SteelBlue, WB-DarkRed, WB-SaddleBrown, WB- Pink, and WB-MediumPurple3. Of these, WB-Tan and WB-DarkRed were also related to sex, while WB-SteelBlue and WB-MediumPurple3 were also related to age. The gene lists for each of these 10 whole blood modules are provided in Supplementary Table 11C.

Genes that are highly interconnected in each time-associated module were also identified for all sample types. These represent “hub” genes with high potential for functional or regulatory significance (Supplemental Table 12). Examples of hub genes from the different modules in monocytes include: *SCAF11*, *CREBZF*, *SETX*, *JAK1*, *EIF3F*, *HNRNPK*, *UBC*, *DICER1*, *CAPN2*, *RAB10*, *SF3B1*, *DAZAP2*, *UTY*, *SPARC*, *PPP4C*, and *RAB11FIP1*. Examples of hub genes from the different modules in whole blood include: *TNFRSF18*, *PCSK9*, *IGSF9*, *MTHFSD*, *DCAF12*, *BCL2L1*, *MAU2*, *TLL2*, *FCRL6*, *PARVG*, and *VASP*.

A tissue-specific network-based functional characterization of the hub genes was performed [36] to gain perspective on the hub genes identified in monocytes (Figure 9B, Supplemental Table 13A) and whole blood (Figure 9C, Supplemental Table 13C). Neutrophils were not analyzed since only two hub genes were found: *RP11-35J10.7* and *AP000593.6* (Supplemental Table 12B).

In monocytes eight functional HumanBase modules were identified and included the terms: response to virus and immune effector process (HG-M1, <u>H</u>ub <u>G</u>enes in <u>M</u>onocytes cluster <u>1</u>), RNA splicing (HG-M2), RNA metabolism (HG-M3), DNA repair (HG-M4), cell morphogenesis (HG-M5), regulation of cell cycle (HG-M3 and HG-M6), cell adhesion and secretion (HG-M7) and protein transport (HG-M8) (Figure 9B and Supplemental Table 13). The terms for HG-M2-4 are unique to monocytes (lists in Supplemental Table 13B). The network-based functional interpretation of the whole blood hub genes in time-related WGCNA modules identified five functional HumanBase modules that included the terms: response to virus and immune effector process (HG- WB1, <u>H</u>ub <u>G</u>enes in <u>W</u>hole <u>B</u>lood cluster <u>1</u>), leukocyte activation (HG-WB2), filopodium assembly (HG-WB3), regulation of cellular response to transforming growth factor beta receptor (HG-WB4), gene silencing (HG-WB4) and cytoskeleton organization (HG-WB5) (Figure 9C and Supplemental Table 13D). The first two terms in HG-WB1 were shared between whole blood and monocytes HG-M1. The rest were only identified in whole blood (overlaps not shown, all lists available in Supplemental Table 13).

There was a significant correlation between the expression of some monocyte and whole blood hub genes with NIHSS on admission (Supplemental Table 14A-B). Examples of hub genes in monocytes that correlate with NIHSS included *MIER1*, *DICER1*, *FBXW5*, *SELPLG*, *TIA1* and *BCKDK* (p < 0.01, Supplemental Table 14A). There is no overlap between the NIHSS significantly-correlated hub genes from monocytes and whole blood (Supplemental Table 14). The hub genes identified in neutrophils included *RP11-35J10.7* which is a novel transcript that is sense intronic to *OVCH2* (ovochymase-2) (Supplemental Table 12B). The second was *AP000593.6*, a U2 small nuclear RNA auxiliary factor 1 (*U2AF1*), currently annotated as a pseudogene (Supplemental Table 12B). Since there were only two neutrophil hub genes, no additional functional analyses were performed for neutrophils.

Gene markers delineating M1/M2 monocyte and N1/N2 neutrophil subsets were identified in the time-related modules. Classical or inflammatory monocyte markers like *CD14*, *CCR2*, *CSF1R* and nonclassical or intermediate marker *FCGR3A* [37, 38] were found in three time-associated modules in monocytes (Supplementary Table 11A) along with M2/N2 polarization-related markers *CCL2* and *STAT3* [8,39–41]. The presence of *CCR2* in a time-related module, the main chemoattractant for monocytes to the injury site in IS [42, 43], may also suggest an active recruitment of this leukocyte type. In neutrophils, none of these markers were present in the modules, while in whole blood *CCR2* and *TNF* (M2 [41, 44] and M1/N1 markers [40,45–47], respectively) were present in two significant time modules (Supplementary Table 11C).

## Discussion

Ischemic stroke elicits specific responses in peripheral blood, which can be seen at the transcriptional level in monocytes and neutrophils [18] as well as other cell types. This is the first study to analyze those differences based on time trajectories in the early time window after stroke using RNA-seq. Hundreds of genes change expression at different time points following stroke, with most genes in neutrophils being up-regulated and most genes in monocytes being down-regulated over time. The genes that change expression at each time point and in each cell type are associated with specific pathways that are usually unique for each cell type and time. Self-organizing maps identified specific trajectories of gene expression for monocytes, neutrophils and whole blood that were similar but were associated with differing signaling pathways. These pathways also differed as a function of cardioembolic, large vessel and small vessel causes of stroke. We also show modules of genes that are co-expressed and change with time following stroke using WGCNA in neutrophils, monocytes and whole blood.

Specific hub genes/ potential master regulators were identified for modules that were associated with time after stroke. These analyses help understand genes and functions changing across the early post-ischemic stroke period and groups of genes that behave in a concerted fashion. Both aspects are important in the search for better understanding of the complex molecular and immune interactions after ischemic stroke to identify optimal treatment targets and their optimal time windows.

### Gene expression dynamics after ischemic stroke

Differential gene expression changes in ischemic stroke (IS) versus controls were assessed by splitting subject samples into different time windows. Monocytes displayed down-regulation of most of their DEGs, while neutrophils were generally up- regulated. This is in accordance with the up- and down-regulation patterns seen in our previous study [18] that did not consider time. For both monocytes and neutrophils, the largest number of differentially expressed genes was observed at the 24-48 h time point, whereas the greatest number of regulated genes in whole blood was observed at the 0-24 h time point. This suggests that additional cell types in whole blood may drive the early peripheral immune response in the first 24 h.

Canonical pathways associated with monocytes over time were mostly suppressed, similar to our previous findings [18]. Many of the over-represented functional pathways are shared in monocytes across time points, with very few shared pathways over time in neutrophils, and none in whole blood. These findings suggest more dynamic changes of genes and pathways in neutrophils and leukocytes, with much less dynamic change in monocytes at least over the first few days after a stroke. When analyzing enrichment of canonical pathways and prediction of upstream regulators, a z-score is calculated with Ingenuity software, where z ≥ 2 is activated (denoted ahead with subscript *act)* and z ≤ −2 is inhibited (denoted ahead with subscript *inh*).

Cytokine expression changes, and specifically those of interleukins, were prominent and some changed with time in monocytes. Since changes of many interleukins have been directly quantified in blood of IS patients over time [48, 49], it was not surprising that we saw many changes of cytokine and interleukin genes representing their receptors. The IL-4 signaling pathway that is essential for M2 polarization [50] was enriched across all time points with IL-4 itself predicted as inhibited based on the observed overall gene expression profile in monocytes at 24-48 h and >48 h in this study. The IL-8 signaling pathway *inh* was also enriched across all time points in monocytes. Pro-inflammatory cytokine IL-8 has a role in monocytic recruitment, by promoting monocyte adhesion to the vascular endothelium [51]. TGF-ꞵ (produced by M2 macrophages) has a protective role in stroke [52] and is inhibited in monocytes only at 24-48 h. IL-6 signaling was enriched and significantly suppressed only at 24-48 h, and IL-15 signaling is suppressed at all time points. IL- 6 (pro- and anti- inflammatory cytokine), is a marker of infarct size and functional outcome, and is part of M2 polarization signaling promoting monocytic differentiation into macrophages [53–59]. IL- 15 is mostly produced by monocytes and its blockade reduces brain injury after ischemia [60, 61]. In monocytes, IL-5*inh*, IL-2*inh* and TNF*inh* were predicted upstream regulators of gene expression at 0-24 h; IL-1B*inh*, IL-2*inh*, IL-3, IL-4*inh*, IL-5*inh*, IL-10*inh*, IL- 13, IL-15, IL-33*inh* and TNF*inh* are predicted upstream regulators at 24-48 h; and IL-2*inh*, IL-3, IL-4*inh*, IL-13, IL-33, and TNF*inh* are upstream regulators at >48 h.

The release of granules from neutrophils results from IL-8 stimulation [62, 63], a pathway that was over-represented only at 24-48 h and predicted as inhibited. Other pathways uniquely enriched between 24-48 h post IS included those for N2 markers TGF-β and IL-4. IL-4 activates neutrophils [64], and along with pro-inflammatory IL-1 and IL-7, are key for T-cell proliferation and homeostasis [65].

The IL-15 pathway *inh* is enriched at 24-48 h and >48 h and IL-15 is identified as an upstream regulator itself at 24-48 h. IL-10 pathway and IL-2 pathway inh were only present in neutrophils at times >48 h. IL-10 is an anti-inflammatory cytokine and N2 marker which decreases inflammation and apoptosis [59]. IL-15 induces changes in neutrophils and increases their phagocytic activity [66]. IL-2*inh*, a T-cell growth factor and activator, is predicted as inhibited in this study which corresponds with the temporal profile for this cytokine in IS [48, 67]. Additionally, the N1 marker GM-CSF signaling pathway, which is protective in experimental stroke [68], was enriched at > 48 h after stroke. IL-2, IL-3, IL-4, IL-5, IL-6, IL-10, IL-11RA, IL-15, IL-16, IL-23A, IL-32 and TNF*inh* were predicted upstream regulators of gene expression in neutrophils at 24-48 h; and IL-2, IL-3, IL-4, IL-5*inh*, IL-10 and TNF*inh* are predicted upstream regulators of gene expression profiles seen in neutrophils at >48 h.

Whole blood over-represented pathways were most robust at 0-24 h after stroke, and rather than being related to cytokines or chemokines, they appear to relate to specific biological responses to ischemia including p53, ATM, cAMP*act* and AMP- activated protein kinase signaling [69–74]. Ephrin receptor signaling *act* that is key in angiogenesis after stroke is over-represented in whole blood at 24-48 h after stroke [75]. In general, fewer canonical pathways were over-represented in whole blood, despite having as many or more DEGs as the monocyte and neutrophil datasets. This could be due to a wide range of biological processes that might be regulated in opposite directions in a heterogeneous sample of multiple cell types from peripheral blood. This highlights the need to further delineate temporal changes of DEGs in each immune cell type following stroke. Though no cytokines were predicted as upstream regulators of gene expression in whole blood, there were a significant number of microRNAs at the three times that were identified as predicted upstream regulators.

The molecules identified as upstream regulators of gene expression in the different cell types might be particularly important therapeutic targets since they have potential to affect so many downstream genes with concerted expression changes after stroke [76]. For example, in neutrophils calcitriol is identified as an upstream regulator at all three time points, but with a trend towards suppression between 0 to 48 h, followed by activation at >48 h after IS. Though calcitriol has shown neuroprotective effects in experimental models of IS [77–79], our human data might point to loss of efficacy outside of an early time window. Pterostilbene, an antioxidant, anti-inflammatory and anti-apoptotic compound with a beneficial effect after stroke [80–83], was predicted as an upstream regulator of the observed changes in neutrophil gene expression but only at times >48h after IS.

### Time-associated gene expression modules and networks in monocytes, neutrophils, and whole blood

By analyzing correlation of co-expression patterns as a continuous variable over time, different modules and their associated hub genes were identified. Hub genes potentially drive the gene expression changes for the different modules of co-expressed genes. The most time-associated modules were associated with monocytes followed by whole blood and then neutrophils.

Monocyte polarization markers are present in four time-associated modules, including the MON-LightCyan module where *CD14* (Classical or inflammatory monocyte marker) and *STAT3* (M2 marker) genes are present. In this module, hub genes are also significantly enriched for monocyte-specific genes, suggesting that the expression dynamics of these genes could be critical for the monocyte response after stroke.

Furthermore, the vast majority of the MON-LightCyan module hub genes display higher expression levels in classical monocytes (The Human Protein Atlas [38]). Other key markers present in time-related modules are *CCR2* (Classical monocyte marker), *CSF1R* (M2 polarization) and *CCL2* (M2 marker). Together, these results may indicate transformation of monocytes to the restorative M2 type over time, even within the ∼72 h time period of this study. Nonetheless, the correlation of *CCR2* with time is consistent with active recruitment of monocytes from the bone marrow, which is in line with experimental data where monocytes begin to accumulate around day 3 or later after ischemic stroke [84].

In neutrophils, the genes in the only time-significant module did not overlap with cell type or polarization markers. In contrast, the gene markers TNF-α (M1 and N1) and *CCR2* (M2) were present in whole blood modules associated with time. As expected, different polarization types are present in whole blood, the same is true for cell type markers, which were significantly enriched in several significant to time modules (monocyte, granulocyte, erythroblast, natural killer and T cell markers). *CCR2* expression in neutrophils is not expected in a resting state, but has been linked to altered neutrophil programming in inflammatory states [85] and promoting chemotactic attraction to the injury site [85].

The functional and biological roles of the highly interconnected time-associated hub genes identified through WGCNA point to unique processes and drivers of gene expression in monocytes and whole blood. Hub genes in monocytes are enriched significantly for RNA splicing and RNA metabolism functions. This may reflect active formation of specifically spliced gene transcripts in the response to injury and timing of polarization which likely changes as monocytes move from inflammatory to restorative subtypes (M1 and M2 monocytes, respectively).

Hub gene functional modules in whole blood represent a composite of the gene expression changes in individual immune cell types. Enrichment of a wide host of functions including leukocyte activation, filopodium assembly, cytoskeleton organization, regulation of cellular response to transforming growth factor beta receptor, and gene silencing point to the breadth of cell types in blood undergoing cell proliferation, activation, and migration in the first 72 h after ischemic stroke.

Several time-associated hub genes were positively correlated with NIHSS (NIH stroke scale, a measure of stroke severity) at admission. In monocytes, a positive significant correlation with NIHSS was found for *TIA1* (T-Cell-Restricted Intracellular Antigen-1 aka cytotoxic granule associated RNA binding protein), an anti-inflammatory protein in peripheral tissues and a repressor of TNF-α expression. It is a M1 marker, and a key regulator of the innate immune response of the CNS during stress [86–88]. *MIER1,* a transcriptional repressor [89], and RMI1, a DNA repair protein [90], were also positively correlated with stroke severity. Variants in these genes are associated with monocyte counts [91], and myeloid white cell counts [91], respectively; and expression positively correlates with the infiltration of monocytes, macrophages and other immune cells in gastric carcinoma [92]. Other NIHSS-time associated hub genes (positive correlation) included: *SELPLG,* high affinity receptor for cell adhesion molecules in leukocytes [93]; *GNAI3*, associated with the response to intracerebral hemorrhage [94] and involved in VEGF-induced angiogenesis [95]; and *FBW5*, an E3 ubiquitin ligase and negative regulator of MAP3K7/TAK1 signaling in the interleukin-1B (IL1B) signaling pathway [96]. IL1B is a key player in the pathogenesis of brain damage after ischemia [97–99] .

In neutrophils, time-associated hub genes did not show a significant correlation with NIHSS at admission. Only one gene in the time-significant module, *MCTS1*, negatively correlated with stroke severity. *MCTS1* is a translation enhancer [100] and is a target of *let7i*, which is involved in leukocyte attachment and recruitment to the endothelium in the brain [101, 102].

In whole blood, immunoglobulin constant region and variable region genes are hub genes in time modules. *IGLV1-40*, IGLV3-27, IGKV1-12, *IGHV3-30* and *IGLC3* showed positive correlation with NIHSS at admission. This highlights changes in the humoral immune response across early times after stroke, and could relate to stroke outcomes [103]. However, given that most of the genes code for variable region chains, this response may also be variable across patients. Further studies are needed to examine these immune humoral profiles and their relationship to evolving stroke injury and repair.

### Refining key genes and responses after IS by analyzing gene clusters GEDI

To generate DEG clusters based on self-organizing maps, tiled mosaics were constructed for each cell type over the three time windows. The GEDI [28] maps allow visualizing coordinated gene expression changes across time for smaller groups of DEGs (1-77 genes per tile/cluster, averaging 11.9 (M), 9.9 (N), 8.4 (WB)), and to visually compare distinct organization of the mosaics between monocytes, neutrophils and whole blood. In the mosaics, tiles of interest are those that have marked differences in expression at a specific time point, those that change consistently through time, and those that maintain constant expression levels across time. From the DEGs grouped in different tiles, monocyte and neutrophil-specific markers were identified. Most tiles containing these markers were neighboring each other, likely because they were correlated by expression through the dimension reduction used to construct the GEDI maps. Nonetheless, in whole blood, the cell-specific marker genes present in some tiles (monocytes, granulocytes, erythroblast, B cells and megakaryocytes) do not group in the same neighborhoods. Analyzing distant cell marker tiles can be critical to refine different functional relevance for the DEGs in those tiles.

In the monocyte GEDI maps, the lower left tile is one case of a cell marker-containing tile that is unrelated to other tiles in the mosaic with other cell-specific markers. This tile/group of DEGs has sustained low expression (namely 1,7) and contains the monocyte marker *CACNA2D4* (Calcium voltage-gated channel auxiliary subunit alpha2delta 4). This gene displays higher expression in classical monocytes [http://www.proteinatlas.org] [38, 104] and belongs to the TCR signaling pathway.

*CACNA2D4* gene expression dynamics, as visible in its GEDI tile, could indicate a switch to non-classical monocytes. Closer examination of the same tile shows other genes including *SURF1* (Cytochrome C Oxidase Assembly Factor) and *TSPAN14*, which are associated with monocyte counts in genetic studies [91]. Other genes in this cluster include *DNPH1* (2’-deoxynucleoside 5’-phosphate N-hydrolase 1), *ZBTB5* (Zinc finger and BTB domain containing 5), *MTBP* (MDM2 binding protein), and six other uncharacterized transcripts, which may share similar roles to the other genes in the tile. These expression correlations still do not fully define shifts towards a cell subtype, like classical, non-classical and intermediate monocytes. For example, *TSPAN14* is highly expressed in non-classical monocytes more than in other subtypes, while *DNPH1* is predominant in intermediate monocytes. Altogether, looking at specific tiles can implicate key genes and cellular processes that change with time after stroke.

In monocytes, “neighborhoods” of tiles with monocytes markers can be seen on the upper and lower parts of the mosaic. These tiles showed different trends across time, from decreasing expression (tile 1,1) to unchanged gene expression (i.e. 1,7; 5,1 and 5,2). The grouping of cell-specific genes is also seen and even more accentuated in neutrophils, where granulocyte markers cluster in four opposite corners of the mosaic, displaying increasing or consistent high expression. Also in these neutrophil mosaics, two upper tiles in opposite corners (namely 2,1 – left and 7,1 – right) rapidly change expression across TPs 1 and 2, and then decrease to reflect levels more like TP0/controls. These tiles also have interleukin receptor genes (Figure 5, depicted by +). Tile 2,8, which contains an immunoglobulin gene (depicted with *) shows a pattern of increasing expression across time. DEGs of interest present in tile 2,8 of the neutrophil mosaic include *TLR5*, an activator of the innate immune response [105], and platelet aggregation gene *PDK1*, key for cell division in hypoxic conditions [106]. This tile also includes relevant genes that have robust expression in healthy granulocytes or neutrophils, but in stroke are more suppressed in TP3, like *KREMEN1*, a negative regulator of Wnt/β catenin pathway [107] and *PPP4R2*, a modulator of neuronal differentiation and survival [108].

Additionally, we consistently see dynamic profiles of immunoglobulin expression in our samples including the whole blood mosaic (left lowermost tiles). These clusters of DEGs peak in the first 24 h and decrease thereafter. Several immunoglobulin genes are expressed as a function of time across samples. B cell activation and increased immunoglobulin production have been demonstrated after stroke [109–111]. Further work could elucidate whether the immunoglobulin genes detected in this study are related to the expected immune response after stroke, or to the production of autoantibodies seen in ischemic stroke patients [112–115]. The latter will be interesting to explore, given the cellular response to brain antigens is associated with infarct size and outcome [116, 117].

### Similar expression dynamics by SOM

Self-Organizing Map (SOM) profiles enable grouping of the DEGs into clusters based on trajectory/directionality of expression changes and their functional associations across the time following stroke. After analyzing GEDI tiles, this is also useful because ontology analyses are more precise in larger gene groups and SOM profiles draw from larger and/or more balanced clusters. These profiles are crucial for understanding which molecular pathways and cell types show progressive activation or suppression over time, or whether they have “peaks” or “valleys” of expression over time.

In monocytes and neutrophils, the profiles of DEGs that peak only in the first 24 h after stroke are enriched for myeloid leukocyte activation and leukocyte migration, respectively. Interleukin-1 beta secretion in monocytes, and positive regulation of reactive oxygen species metabolism in neutrophils, are overrepresented in profiles that decrease expression over time. Genes that are suppressed between 0-24 h and then recover to levels seen in VRFCs, are enriched for terms related to JNK and MAPK cascades, platelet activation and macrophage chemotaxis in monocytes. This pattern also has over-representation of chemokine production and leukocyte aggregation genes in neutrophils. Opposite trends can also be seen between monocytes and neutrophils: Cdc42-associated signaling (critical in cell growth and differentiation [118]) is enriched in DEGs that decrease expression over time in monocytes; while in neutrophils Cdc42- associated signaling is associated with genes that increase expression over time.

Furthermore, the trajectory of the SOM profiles detected in the DEGs at different time points could be used to prioritize diagnostic biomarker candidates. A diagnostic panel that could be used to diagnose stroke in the first 3 days should primarily include DEGs that either consistently increase or consistently decrease over the 3 days after IS. Such panels could indicate not only that an ischemic stroke had occurred, but could indicate roughly how much time had elapsed following the stroke – something that cannot be estimated with current methodology.

### Refining genes for cardioembolic, large vessel, and small vessel strokes

There were large numbers of DEGs expressed in monocytes and neutrophils for each cause of stroke (CE, LV, SV) with somewhat more DEGs 24 h after stroke. In contrast there were many more DEGs expressed in whole blood at 0-24 h in CE, LV and SV stroke. This suggests that other cells (e.g., B and T cells) in whole blood (in addition to neutrophils and monocytes) were contributing to the whole blood responses to stroke at 0-24 h.

The trajectories of SOM profiles in strokes of all causes combined were similar to profiles for each stroke cause including CE, LV and SV causes of strokes. Though the temporal profiles were similar, the DEGs and enriched GO terms were generally different for CE, LV and SV causes of stroke in monocytes, neutrophils and whole blood. There were exceptions, such as the “Regulation of Golgi to plasma membrane protein transport” pathway, which progressively decreased in expression in monocytes in CE, LV and SV stroke, a pathway shown to affect outcomes in experimental stroke [119]. Another example was “NADH oxidation” which peaked at 24 h in neutrophils in CE, LV and SV strokes and is known to play a role in experimental stroke [120]. There were few shared GO terms enriched in DEGs in whole blood CE, LV and SV strokes, likely related to the cellular heterogeneity of whole blood.

SOM profiles identified important attributes of DEGs from opposite expression trajectories and between IS etiologies. In neutrophils, the toll-like receptor signaling pathway is associated with a profile that decreases at less than 24 h and increases after 24 h in CE stroke. In contrast, for LV, the toll-like receptor pathway is enriched in a profile that decreases at all times compared to controls. Toll-like receptor signaling modulates critical immunomodulatory NFkB signaling and is a promising target for treating cardiovascular disease [121–123] (clinicaltrials.gov ID# <u>NCT04734548</u>). TLR signaling impacts downstream pro- or anti-inflammatory molecules, including TNFα, interleukins, interferons and TGF-β [124].

There are also differences between SOM profiles in whole blood for different causes of stroke. “Positive regulation of IL-1β secretion” genes peak in the first 24 h in CE and LV strokes, whereas in SV stroke IL-1 genes decrease at 24 h and then recover at times after 24 h. An example of the complexity of the profiles is observed for whole blood in SV strokes where both a peaking of positive regulation of cytokines is noted at 24 h with decreases thereafter, as well as consistent increase over time of pathways associated with negative regulation of cytokines. Thus, either different cell types in whole blood are responding differently, and/or different cytokines are being regulated differently. This serves to emphasize how complex the temporal and cellular responses are to stroke, and to warn against time-agnostic approaches to treatment at single times of strokes of different causes.

### Limitations

Though it was possible to identify markers for monocyte and neutrophil polarization in either direction (inflammatory or anti-inflammatory types), it was not possible to define a unique shift towards a specific subtype since we expect that both M1/M2 and N1/N2 phenotypes are present in the samples of pooled cells in the ∼3 day time window studied here. It is possible that studying longer times after stroke might allow detection of these shifts in gene expression responsible for the evolution into polarized M2 or N2 phenotypes. Moreover, future single cell RNA-seq studies should enable a better identification of the different cell subpopulations present at each time point.

This study employed single samples from single patients at different times. Thus, the changes of gene expression represent the average of multiple patients over the times stipulated. We have shown that individual genetic variation plays a key role in the specific expression response after stroke [125]. Our approach presented here based on the availability of samples would include inter-individual variability of expression as compared to differences measured in a single subject over time. Thus, future studies should consider a serial sample approach in every subject over longer periods of time to investigate gene expression dynamics in every subject and see how these vary between subjects as a function of time and its relation to infarct volume, evolving NIHSS, and other clinical variables.

A strength of the study was the multiple analytical approaches used, including differential expression, GEDI, SOM and WGCNA, with each providing insight to the complexity and dynamics of gene expression changes after stroke. SOM in particular was able to demonstrate how pathways changed over time for each cell type and cause of stroke. WGCNA identified modules of co-expressed genes and associated hub genes that changed over time. However, even WGCNA had some weaknesses in that few hub genes and only one time-associated module were identified in neutrophils. Both these results likely arose from differences in size of input list of genes or specific parameters used in the models, since we indeed observed slightly more time-regulated DEGs in neutrophils compared to monocytes when studied by time point. Since WGCNA identified modules that correlated with the continuous time parameter, it may have missed modules with complex dynamic profiles (like the identified SOM profiles).

Pathway and functional analyses helped to interpret aggregate shifts of groups of genes, while reducing the impact of potential false positives of individual genes. Though these results are likely to be more reliable, they are hampered in the current study by the small sample size for many of the time points particularly for those considering different causes of stroke in the three sample types examined (isolated monocytes, isolated neutrophils, whole peripheral blood). Much larger sample sizes of blood samples collected over multiple times in each subject will be needed for future studies. Overall, however, each of these analytical approaches provided unique insights into the pathophysiology of stroke based upon cells in blood which modulate the peripheral clotting and immune responses in stroke.

## Conclusions

- We identified key genes associated with time in the response from leukocytes after ischemic stroke. Some of these correlated with stroke severity.
- Differentially expressed genes have distinctive trajectories for all IS etiologies analyzed, which allows refinement of critical genes and functions at specific time points after stroke.
- Altogether, the identified changes in gene expression and pathways over time are critical for understanding how the immune and clotting systems change dynamically after stroke, and point to the complexities of identifying biomarkers and treatment targets for stroke.

## Data Availability

The supplementary material is available online as indicated below. Other data used and/or analyzed during the current study are available from the corresponding author on reasonable request.

## Abbreviations

IS: Ischemic Stroke
VRFC: Vascular risk factor control
NIHSS: National Institutes of Health stroke scale
WGCNA: Weighted Gene Co-expression Network Analysis
GO: Gene ontology
DEGs: Differentially expressed genes
DE: Differentially expressed
TP: Time point
GEDI: Gene expression dynamics inspector
SOM: Self-organizing maps
CE: cardioembolic
LV: Large vessel
SV: Small vessel
IL: Interleukin
ATM: ataxia-telangiectasia mutated ATM kinase

## Declarations

### Ethics approval and consent to participate

This study was reviewed and approved by the University of California, Davis Institutional Review Board (IRB ID# 248994-41). All participants or a legally authorized representative provided written informed consent to participate in this study.

### Consent for publication

Not applicable

### Authors’ contributions

Conceived and designed the experiments, and edited the manuscript: PC-M, BPA, BS, FRS. Performed the experiments and/or reviewed data: PC-M, BK, MH, HH, XZ, NA, HA. Clinical characterization: GCJ, FRS. Wrote the first draft: PC-M.

## Acknowledgements

We thank the patients and families that participated in the study. The sequencing was carried out by the DNA Technologies and Expression Analysis Core at the UC Davis Genome Center, supported by NIH Shared Instrumentation Grant 1S10OD010786-01. Monocyte and neutrophil clip art from Servier https://smart.servier.com/ and licensed under CC-BY 3.0 Unported https://creativecommons.org/licenses/by/3.0/.

## Supplemental Figures

**Supplemental Figure 1. Percentage of DEGs classified per biotype.** For every time point and comparison between time points, the percentage of biotype categories for DEGs are shown in the bar plots and corresponding table for (A) monocytes, (B) neutrophils and (C) whole blood. Ig: immunoglobulin; TcR: T-cell receptor; TEC: To be Experimentally Confirmed.

**Supplemental Figure 2. Predicted upstream regulators for monocytes.** (A) Venn diagram represents all upstream regulators predicted at 0-24 h, 24-48 h, and >48 h (Fisher’s p-value<0.05). (B) Top over-represented regulators common to all time points (Fisher’s p-value<0.05). (C) Top over-represented regulators that are specific for each time point (Fisher’s p-value<0.05). Up arrows indicate predicted significant activation (z ≥ 2) or down arrow - significant inhibition (z ≤ −2). White cells indicate no direction can be predicted. (*= Benjamini-Hochberg corrected p-value<0.05). R: receptor; N.R: nuclear receptor.

**Supplemental Figure 3. Predicted upstream regulators for neutrophils.** (A) Venn diagram represents all upstream regulators predicted at 0-24 h, 24-48 h, and >48 h (Fisher’s p-value<0.05). (B) Top over-represented regulators shared at all time points (Fisher’s p-value<0.05). (C) Top over-represented regulators that are specific for each time point (Fisher’s p-value<0.05). Up arrows indicate predicted significant activation (z ≥ 2) or down arrow - significant inhibition (z ≤ −2). White cells indicate no direction can be predicted. (*= Benjamini-Hochberg corrected p-value<0.05). R: receptor; endo: endogenous; reg: regulator.

**Supplemental Figure 4. Predicted upstream regulators for whole blood.** (A) Venn diagram represents all upstream regulators predicted at 0-24 h, 24-48 h, and >48 h (Fisher’s p-value<0.05). (B) Only 2 predicted regulators (Fisher’s p-value<0.05) are shared for the 3 time points. (C) Top over-represented regulators that are specific for each time point (Fisher’s p-value<0.05). Up arrows indicate predicted significant activation (z ≥ 2) or down arrow - significant inhibition (z ≤ −2). White cells indicate no direction can be predicted. R: receptor; reg: regulator; endo: endogenous.

**Supplemental Figure 5. Co-expression network construction.** WGCNA Soft-thresholding power scale free topology fit (left panel) and mean connectivity (right panel) plots for all genes analyzed in (A) monocytes, (B) neutrophils and (C) whole blood. Reference lines are at 0.8 (red) and 0.9 (green) on the left panels and at 100 (red) and 200 (green) on the right panels.

