## Supplemental figures for "Monocyte, Neutrophil and Whole Blood Transcriptome Dynamics Following Ischemic Stroke"

### Sup. Figure 1

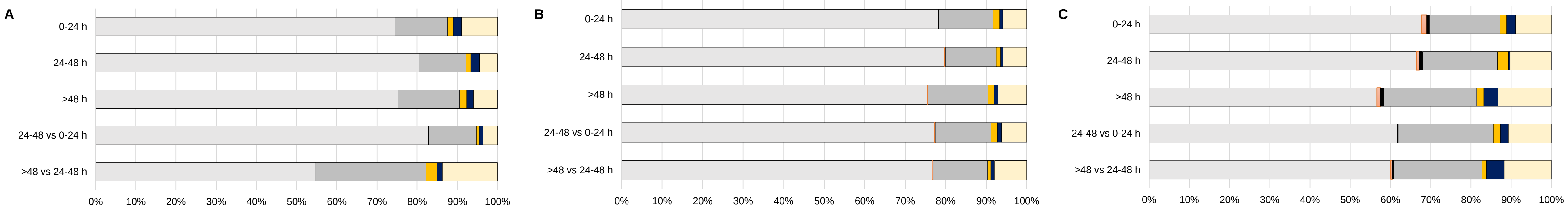


| Monocytes | >48 vs 24-48 h | 24-48 vs 0-24 h | >48 h | 24-48 h | 0-24 h |
|---|---|---|---|---|---|
| Protein Coding | 54.8 | 82.7 | 75.0 | 80.5 | 74.5 |
| Ig | 0.0 | 0.0 | 0.0 | 0.0 | 0.0 |
| TcR | 0.0 | 0.3 | 0.0 | 0.0 | 0.0 |
| lncRNA | 27.4 | 11.8 | 15.3 | 11.6 | 13.1 |
| Non-coding RNA | 2.7 | 0.7 | 1.7 | 1.2 | 1.4 |
| TEC | 1.4 | 0.9 | 1.7 | 2.2 | 2.1 |
| Pseudogene | 13.7 | 3.6 | 6.0 | 4.5 | 9.0 |

| Neutrophils | >48 vs 24-48 h | 24-48 vs 0-24 h | >48 h | 24-48 h | 0-24 h |
|---|---|---|---|---|---|
| Protein Coding | 76.7 | 77.1 | 75.5 | 78.7 | 78.1 |
| Ig | 0.2 | 0.2 | 0.2 | 0.1 | 0.0 |
| TcR | 0.0 | 0.0 | 0.0 | 0.1 | 0.2 |
| lncRNA | 13.5 | 13.7 | 14.8 | 12.3 | 13.4 |
| Non-coding RNA | 0.7 | 1.6 | 1.5 | 1.1 | 1.6 |
| TEC | 1.0 | 1.1 | 0.9 | 0.5 | 0.8 |
| Pseudogene | 7.9 | 6.2 | 7.1 | 5.8 | 5.9 |

| Whole Blood | >48 vs 24-48 h | 24-48 vs 0-24 h | >48 h | 24-48 h | 0-24 h |
|---|---|---|---|---|---|
| Protein Coding | 60.1 | 61.7 | 56.4 | 64.6 | 67.7 |
| Ig | 0.4 | 0.0 | 0.9 | 0.8 | 1.3 |
| TcR | 0.4 | 0.3 | 0.9 | 0.8 | 0.7 |
| lncRNA | 22.0 | 23.6 | 22.9 | 18.1 | 17.5 |
| Non-coding RNA | 1.1 | 1.8 | 1.8 | 2.7 | 1.6 |
| TEC | 4.4 | 2.1 | 3.5 | 0.4 | 2.3 |
| Pseudogene | 11.7 | 10.6 | 13.2 | 10.0 | 8.9 |

### Sup. Figure 2

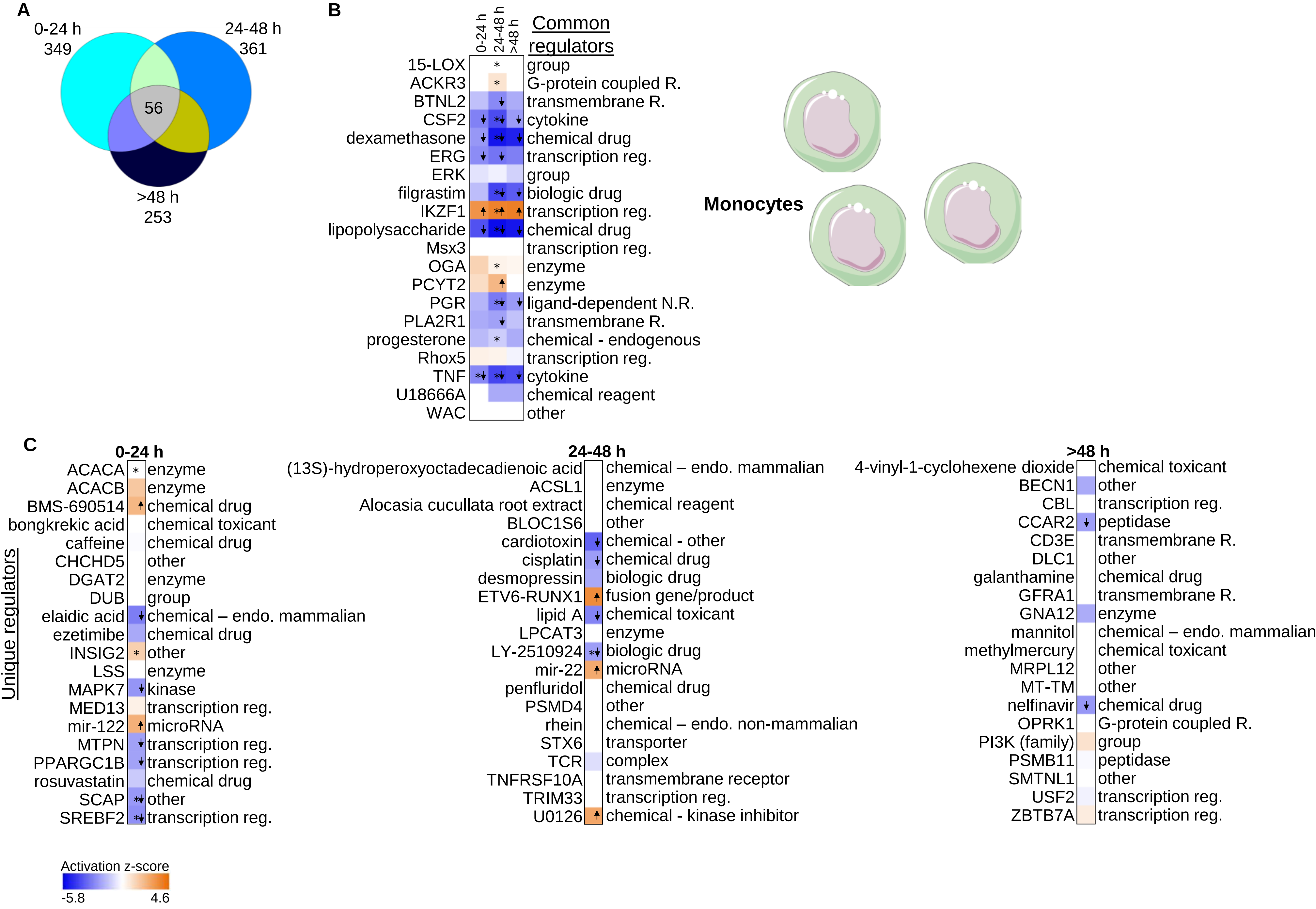
A
0-24 h
349
24-48 h
361
56
>48 h
253
B
0-24 h
24-48 h
>48 h
Common regulators
15-LOX group
ACKR3 G-protein coupled R.
BTNL2 transmembrane R.
CSF2 cytokine
dexamethasone chemical drug
ERG transcription reg.
ERK group
filgrastim biologic drug
IKZF1 transcription reg.
lipopolysaccharide chemical drug
Msx3 transcription reg.
OGA enzyme
PCYT2 enzyme
PGR ligand-dependent N.R.
PLA2R1 transmembrane R.
progesterone chemical - endogenous
Rhox5 transcription reg.
TNF cytokine
U18666A chemical reagent
WAC other
Monocytes
C
Unique regulators
0-24 h
ACACA enzyme
ACACB enzyme
BMS-690514 chemical drug
bongkrekic acid chemical toxicant
caffeine chemical drug
CHCHD5 other
DGAT2 enzyme
DUB group
elaidic acid chemical – endo. mammalian
ezetimibe chemical drug
INSIG2 other
LSS enzyme
MAPK7 kinase
MED13 transcription reg.
mir-122 microRNA
MTPN transcription reg.
PPARGC1B transcription reg.
rosuvastatin chemical drug
SCAP other
SREBF2 transcription reg.
24-48 h
(13S)-hydroperoxyoctadecadienoic acid chemical – endo. mammalian
ACSL1 enzyme
Alocasia cucullata root extract chemical reagent
BLOC1S6 other
cardiotoxin chemical - other
cisplatin chemical drug
desmopressin biologic drug
ETV6-RUNX1 fusion gene/product
lipid A chemical toxicant
LPCAT3 enzyme
LY-2510924 biologic drug
mir-22 microRNA
penfluridol chemical drug
PSMD4 other
rhein chemical – endo. non-mammalian
STX6 transporter
TCR complex
TNFRSF10A transmembrane receptor
TRIM33 transcription reg.
U0126 chemical - kinase inhibitor
>48 h
4-vinyl-1-cyclohexene dioxide chemical toxicant
BECN1 other
CBL transcription reg.
CCAR2 peptidase
CD3E transmembrane R.
DLC1 other
galanthamine chemical drug
GFRA1 transmembrane R.
GNA12 enzyme
mannitol chemical – endo. mammalian
methylmercury chemical toxicant
MRPL12 other
MT-TM other
nelfinavir chemical drug
OPRK1 G-protein coupled R.
PI3K (family) group
PSMB11 peptidase
SMTNL1 other
USF2 transcription reg.
ZBTB7A transcription reg.
Activation z-score
-5.8
4.6

### Sup. Figure 3

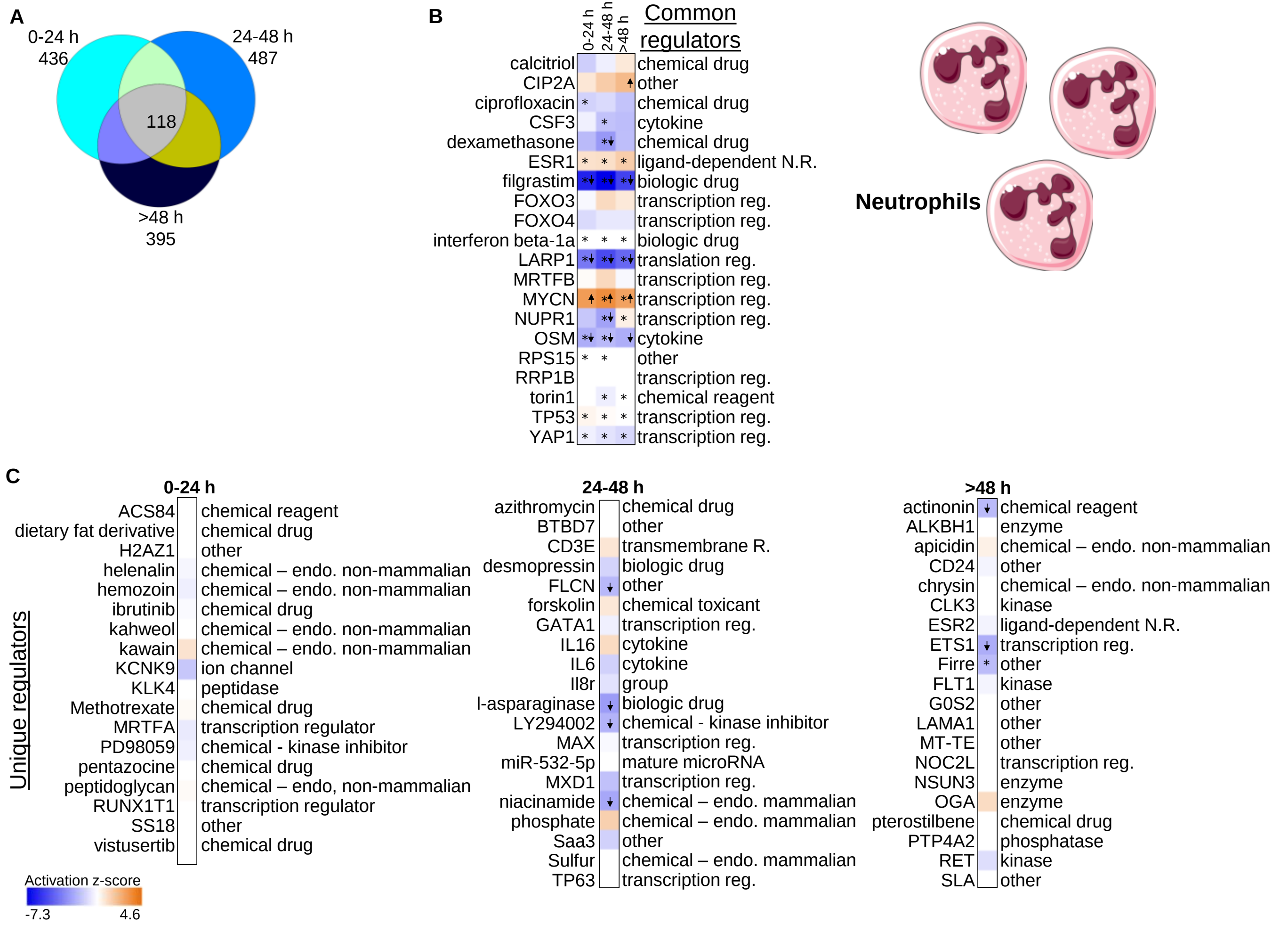

A
0-24 h
436
24-48 h
487
118
>48 h
395
B
0-24 h
24-48 h
>48 h
Common regulators
calcitriol chemical drug
CIP2A other
ciprofloxacin chemical drug
CSF3 cytokine
dexamethasone chemical drug
ESR1 ligand-dependent N.R.
filgrastim biologic drug
FOXO3 transcription reg.
FOXO4 transcription reg.
interferon beta-1a biologic drug
LARP1 translation reg.
MRTFB transcription reg.
MYCN transcription reg.
NUPR1 transcription reg.
OSM cytokine
RPS15 other
RRP1B transcription reg.
torin1 chemical reagent
TP53 transcription reg.
YAP1 transcription reg.
Neutrophils
C
Unique regulators
0-24 h
ACS84 chemical reagent
dietary fat derivative chemical drug
H2AZ1 other
helenalin chemical – endo. non-mammalian
hemozoin chemical – endo. non-mammalian
ibrutinib chemical drug
kahweol chemical – endo. non-mammalian
kawain chemical – endo. non-mammalian
KCNK9 ion channel
KLK4 peptidase
Methotrexate chemical drug
MRTFA transcription regulator
PD98059 chemical - kinase inhibitor
pentazocine chemical drug
peptidoglycan chemical – endo, non-mammalian
RUNX1T1 transcription regulator
SS18 other
vistusertib chemical drug
24-48 h
azithromycin chemical drug
BTBD7 other
CD3E transmembrane R.
desmopressin biologic drug
FLCN other
forskolin chemical toxicant
GATA1 transcription reg.
IL16 cytokine
IL6 cytokine
Il8r group
l-asparaginase biologic drug
LY294002 chemical - kinase inhibitor
MAX transcription reg.
miR-532-5p mature microRNA
MXD1 transcription reg.
niacinamide chemical – endo. mammalian
phosphate chemical – endo. mammalian
Saa3 other
Sulfur chemical – endo. mammalian
TP63 transcription reg.
>48 h
actinonin chemical reagent
ALKBH1 enzyme
apicidin chemical – endo. non-mammalian
CD24 other
chrysin chemical – endo. non-mammalian
CLK3 kinase
ESR2 ligand-dependent N.R.
ETS1 transcription reg.
Firre other
FLT1 kinase
G0S2 other
LAMA1 other
MT-TE other
NOC2L transcription reg.
NSUN3 enzyme
OGA enzyme
pterostilbene chemical drug
PTP4A2 phosphatase
RET kinase
SLA other
Activation z-score
-7.3
4.6

**Sup. Figure 4**

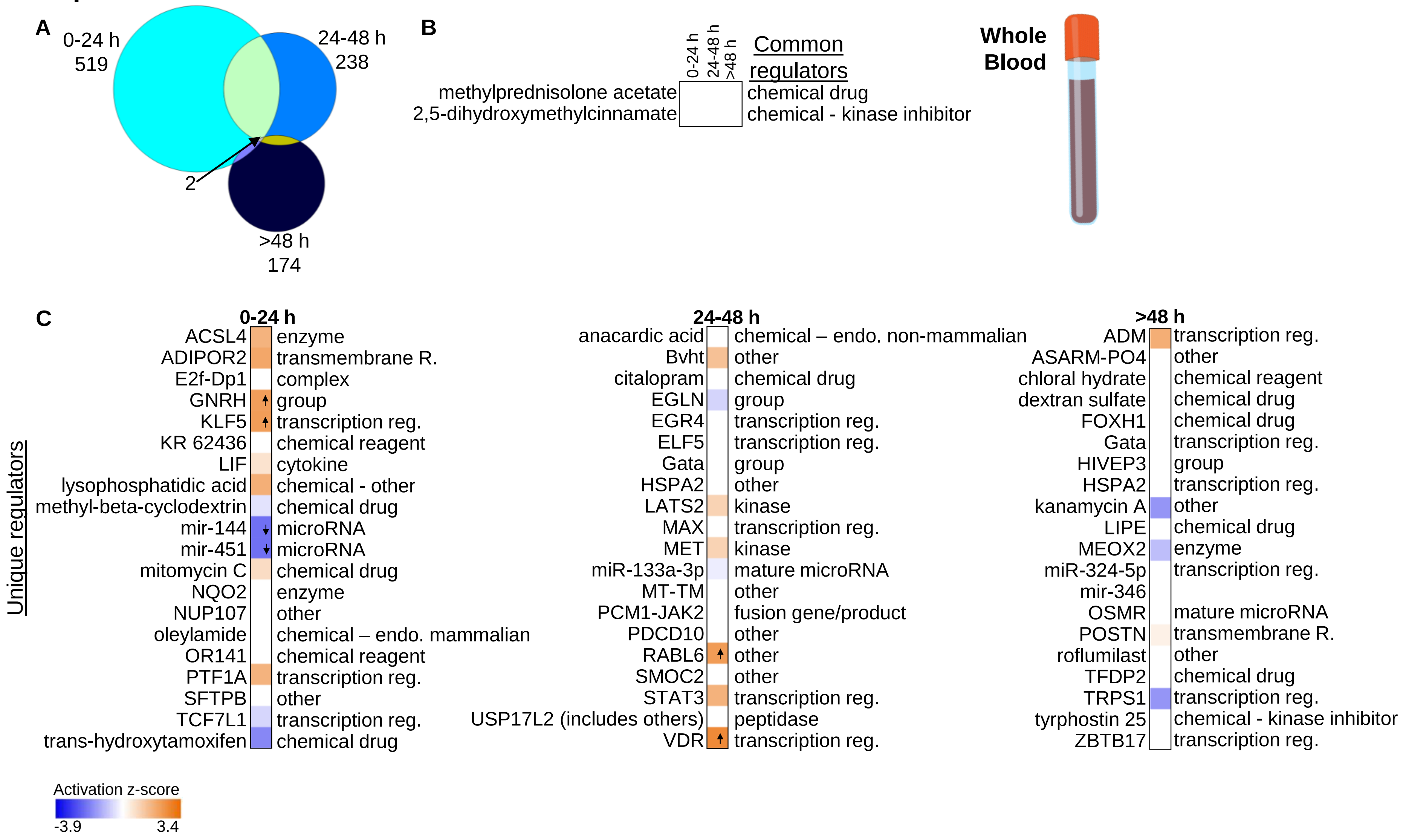
A
0-24 h
519
24-48 h
238
2
>48 h
174
B
0-24 h
24-48 h
>48 h
Common regulators
methylprednisolone acetate
2,5-dihydroxymethylcinnamate
chemical drug
chemical - kinase inhibitor
Whole Blood
C
Unique regulators
0-24 h
ACSL4 enzyme
ADIPOR2 transmembrane R.
E2f-Dp1 complex
GNRH group
KLF5 transcription reg.
KR 62436 chemical reagent
LIF cytokine
lysophosphatidic acid chemical - other
methyl-beta-cyclodextrin chemical drug
mir-144 microRNA
mir-451 microRNA
mitomycin C chemical drug
NQO2 enzyme
NUP107 other
oleylamide chemical – endo. mammalian
OR141 chemical reagent
PTF1A transcription reg.
SFTPB other
TCF7L1 transcription reg.
trans-hydroxytamoxifen chemical drug
24-48 h
anacardic acid chemical – endo. non-mammalian
Bvht other
citalopram chemical drug
EGLN group
EGR4 transcription reg.
ELF5 transcription reg.
Gata group
HSPA2 other
LATS2 kinase
MAX transcription reg.
MET kinase
miR-133a-3p mature microRNA
MT-TM other
PCM1-JAK2 fusion gene/product
PDCD10 other
RABL6 other
SMOC2 other
STAT3 transcription reg.
USP17L2 (includes others) peptidase
VDR transcription reg.
>48 h
ADM transcription reg.
ASARM-PO4 other
chloral hydrate chemical reagent
dextran sulfate chemical drug
FOXH1 chemical drug
Gata transcription reg.
HIVEP3 group
HSPA2 transcription reg.
kanamycin A other
LIPE chemical drug
MEOX2 enzyme
miR-324-5p transcription reg.
mir-346
OSMR mature microRNA
POSTN transmembrane R.
roflumilast other
TFDP2 chemical drug
TRPS1 transcription reg.
tyrphostin 25 chemical - kinase inhibitor
ZBTB17 transcription reg.
Activation z-score
-3.9
3.4

Sup. Figure 5

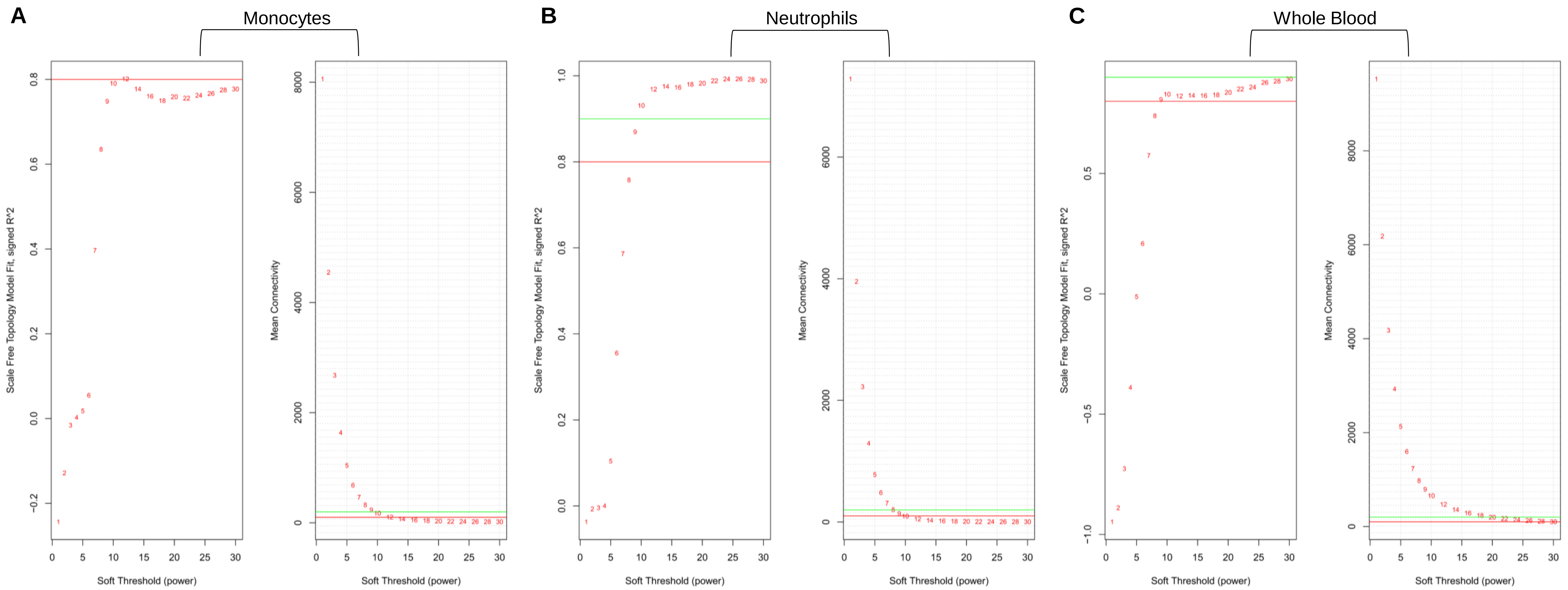

A
Monocytes
B
Neutrophils
C
Whole Blood
Scale Free Topology Model Fit, signed R^2
Mean Connectivity
Soft Threshold (power)
